# Transcriptional Network Dysregulation in Alzheimer’s Disease Revealed by Individual-Specific Gene Regulatory Models

**DOI:** 10.1101/2025.03.26.25324703

**Authors:** Danish Anwer, Eduard Kerkhoven, Annikka Polster

## Abstract

Alzheimer’s disease (AD) is a multifactorial neurodegenerative disorder marked by progressive cognitive decline, yet its transcriptional regulatory architecture remains poorly understood. Here, we model sample-specific gene regulatory networks (GRNs) from dorsolateral prefrontal cortex transcriptomes of 87 individuals with AD and 67 non-cognitively impaired (NCI) controls using PANDA and LIONESS algorithms. This personalized network approach captures individual-level transcriptional regulation, revealing 22 key transcription factor–gene interactions that distinguish AD from NCI with 96% accuracy.

Network topology analyses uncovered significant structural differences in regulatory connectivity and modular organization between AD and controls. Notably, ZNF225, ZNF849, and ZNF548 emerged as AD-specific regulatory hubs. The key transcription factor–gene interactions were enriched in pathways related to ADs molecular presentation, including synaptic signaling, mitochondrial function, proteostasis, and neuroinflammation.

Our findings highlight pervasive transcriptional network dysregulation in AD and underscore the value of personalized GRN modeling in uncovering regulatory mechanisms not apparent from differential gene expression alone. This systems-level approach reveals potential biomarkers and therapeutic targets, offering new insights into the molecular basis of AD and informing precision aging research.

## Introduction

Alzheimer’s disease (AD) is the most prevalent form of dementia, characterized by progressive cognitive decline, memory impairment, and eventual loss of autonomy and death. With over 50 million people currently affected worldwide, and prevalence rising due to increasing life expectancy^1^, AD represents a significant global health burden. In addition to its personal and societal costs, the disease remains without a cure or effective preventive intervention, underscoring the urgent need to understand its underlying molecular mechanisms^2^.

Although the hallmark pathological features, amyloid-β plaques and neurofibrillary tangles, have been studied for decades^3^, the mechanisms underlying AD pathophysiology and progression remain incompletely understood. Genome-wide association studies (GWAS) have identified over 400 AD-associated loci^4^, yet these variants typically exhibit small effect sizes and collectively account for only a modest portion of heritability^5^. This gap, often referred to as “missing heritability”, suggests that complex, systems-level interactions between genes may play a larger role in AD pathogenesis than previously appreciated^6^. Recent conceptual frameworks, such as the omnigenic model^7^, propose that the functional impact of disease-associated variation is distributed across vast gene regulatory networks (GRNs), in which both core and peripheral genes contribute to disease through coordinated transcriptional regulation. In this view, transcriptional dysregulation in disease-relevant tissues, in this case the brain, is likely mediated not by isolated genes, but by disruptions in tissue-specific regulatory networks that govern key cellular processes.

GRN-based approaches thus offer a promising systems-level lens through which to investigate AD. Unlike differential expression analyses that identify gene-level changes, GRNs provide insight into regulatory logic: how transcription factors (TFs) control gene expression and how those interactions change in disease^7^. Prior studies have demonstrated the relevance of GRNs in neurodegenerative disorders, but most analyses to date rely on aggregated population-level data^8,9^. Such approaches overlook inter-individual variation, a critical limitation in AD, where clinical and molecular heterogeneity is important.

In this study, we apply a personalized systems biology approach to model sample-specific gene regulatory networks from postmortem dorsolateral prefrontal cortex (DLPFC) transcriptomes of individuals with and without AD. Using PANDA^10^ and LIONESS^11^ algorithms, we reconstruct individual TF–target gene networks for each study participant and analyse them with machine learning feature selection to identify a minimal yet informative set of regulatory interactions that distinguish AD from non-cognitively impaired (NCI) individuals. By focusing on transcriptional regulation rather than individual gene expression alone, we aim to reveal clinically relevant network-level mechanisms that underpin disease pathophysiology.

Taken together, we aim to define core regulatory modules and transcriptional hubs that are consistently perturbed in AD, providing a systems-level understanding of disease mechanisms and a foundation for precision therapeutics.

## Methods

### Data Acquisition and Cohort Selection

Transcriptomic data from the dorsolateral prefrontal cortex (DLPFC) were obtained from the Religious Orders Study and Memory and Aging Project (ROSMAP)^12^, a longitudinal cohort that integrates detailed antemortem cognitive assessments with postmortem neuropathological evaluations to study aging and neurodegeneration.

Cognitive status was determined using the Cogdx consensus diagnosis^14^, based on structured clinical assessments conducted by expert neurologists. Neuropathological classification followed the Consortium to Establish a Registry for Alzheimer’s Disease (CERAD) criteria^15^, which assign AD pathology levels based on semiquantitative assessment of neuritic plaque burden.

To ensure diagnostic specificity and minimize potential confounding from mixed pathologies or misclassification, we jointly applied both Cogdx and CERAD criteria for cohort selection. Only individuals meeting CERAD criteria for Definite AD and classified as Probable AD by Cogdx, without alternative causes of cognitive impairment, were included as AD cases. Non-cognitively impaired (NCI) controls were defined as those without evidence of AD by either clinical (Cogdx) or neuropathological (CERAD) evaluation.

This combined selection strategy ensured a well-defined case–control structure and reduced the influence of comorbid conditions on transcriptomic analyses.

### Data preprocessing

To retain only biologically relevant genes for analysis, we applied a count-per-million (CPM) threshold to remove low-expressed genes, excluding those with fewer than one CPM in more than 50% of both AD and NCI samples.

To assess and correct for potential batch effects, we conducted an ANOVA test on principal component analysis (PCA) components to determine whether batch-associated variability was significant. To address the identified batch effects, we applied Combat-Seq^16^ for batch correction. However, batch effects remained significant even after correction, thus we excluded batches with persistent effects on gene expression despite Combat-Seq correction.

Additionally, to refine RNA-Seq expression data and minimize the influence of extreme values, we applied quantile-based filtering (5th and 95th percentiles) on PCA components, removing samples with outlier expression patterns. This approach improved dataset integrity by reducing the impact of aberrant values that could skew downstream analyses.

Finally, to address technical biases and ensure comparability across samples, we implemented Median Ratio Normalization (MRN) on raw counts.

### Gene Regulatory Network modelling

To construct aggregate gene regulatory networks (GRNs) as basis to infer individual-specific GRNs for Alzheimer’s disease (AD) and no cognitive impairment (NCI), we employed PANDA (Passing Attributes between Networks for Data Assimilation)^10^. PANDA integrates multiple data modalities (gene expression data, a prior transcription factor (TF)-gene interaction network, and protein-protein interactions (PPIs), to infer comprehensive regulatory networks (Figure 1).

**Figure 1.**
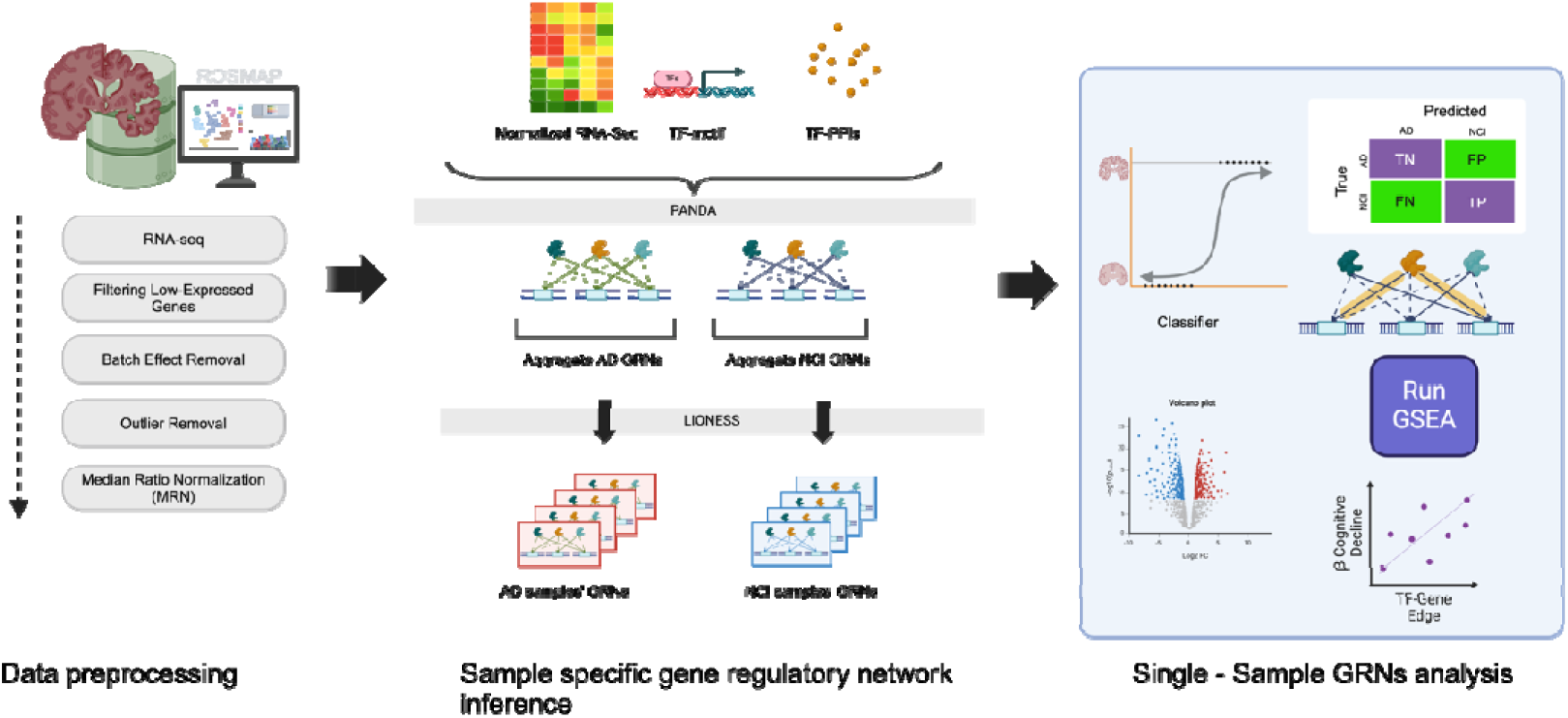
Study Overview. The figure presents the stepwise workflow of the integrative analysis pipeline. Step 1 involves data preprocessing, where quality control, normalization, and transformation techniques are applied to ensure data consistency and reliability. Step 2 focuses on constructing sample-specific gene regulatory networks (GRNs) to capture transcription factor (TF)-gene interactions unique to each sample or condition. Step 3 involves identifying key TF-gene regulatory edges based on network topology and statistical significance. Gene set enrichment analysis is then conducted to reveal biologically relevant pathways and functional categories associated with the identified regulatory interactions. Additionally, correlation analysis between TF-gene interactions and cognitive decline measures, along with differential expression analysis, is performed to gain deeper insights into the underlying molecular mechanisms.

For this study, PPIs were sourced from STRING^17^ with a confidence threshold of 0.5, while TF-motif interactions were obtained from JASPAR^18^. The resulting networks incorporate a regulatory strength metric corresponding to edge weights, quantifying the influence of a TF on its target genes by integrating evidence from expression data, TF-PPI relationships, and TF-motif associations. This metric provides a quantitative measure of transcriptional regulation, reflecting cumulative regulatory impact across data sources.

To assess inter-individual variability in transcriptional regulation, we applied LIONESS^11^ to reconstruct individual-specific GRNs. This approach estimates individual TF-gene interactions, with edge weights reflecting transcriptional regulation strength (Figure 1).

To account for disease-specific regulatory patterns, we derived AD-specific GRNs from the AD aggregate network and NCI-specific GRNs from the NCI aggregate network. By constructing these personalized regulatory networks, we aimed to capture inter-individual heterogeneity, reflecting individual differences in transcriptional regulation that may underly individual disease characteristics, disease susceptibility and trajectory.

To enhance the sensitivity of downstream analyses and improve the biological relevance of detected regulatory mechanisms, we focused on the top 5% of edge weights for common regulatory interactions across GRNs, resulting in a threshold of 3.67 for NCI and 3.71 for AD (Cumulative distribution of edge weights for both AD and NCI are shown in supplementary Figure S1). This stringent filtering approach ensured the identification of the most robust regulatory interactions, facilitating a precise characterization of key transcriptional mechanisms distinguishing AD from NCI.

### Network Analysis of AD-Specific and NCI-Specific GRNs

To analyse the structural properties and identify potential differences in AD-specific and GRN-specific GRNs, we calculated key network metrics for both AD-specific and NCI-specific GRNs. The metrics included density, which measures the proportion of actual connections to possible connections in the network; average degree, representing the average number of connections per node; modularity, quantifying the extent to which the network can be divided into distinct communities; largest component size, indicating the size of the largest connected subgraph; and nestedness, assessing the hierarchical structure of the network. These metrics were calculated to provide insights into the connectivity, robustness, and organization of the regulatory networks, helping to reveal how TF-gene interactions differ in the context of AD and NCI based on network structure.

### Identification of Differentially Regulated Transcription Factor-Target Gene Pairs

To identify differentially regulated target genes and their associated transcription factors (TFs), we employed logistic regression^19^ to classify AD and NCI samples based on edge weights within gene regulatory networks (GRNs). To refine this classification, we applied Recursive Feature Elimination (RFE)^20^, systematically selecting key TFs and target genes most critical for distinguishing AD from NCI.

To assess the statistical significance of differentially regulated edges, we performed a bootstrapping analysis, randomly selecting 22, 100, 500, and 1000 TF-target gene pairs and conducting 1000 iterations for each subset. For each iteration, we calculated classification accuracy, ensuring that the identified regulatory interactions were robust and non-random.

To further validate our findings, we implemented 5-fold cross-validation, assessing the generalizability and stability of the classification model across different data subsets. This approach ensured that our results were reliable and reproducible, strengthening confidence in the detected regulatory mechanisms underlying AD pathophysiology.

### Pathway and Cell Type-Specific Enrichment Analysis

To investigate the functional relevance of key transcription factors (TFs) and their target genes within gene regulatory networks (GRNs), we performed Gene Set Enrichment Analysis (GSEA)^21^, incorporating each TF’s immediate network neighbors. GSEA was conducted using the GSEAPY Python package^22^, with gene sets sourced from the GO Molecular Function 2023, GO Cellular Component 2023, KEGG 2021 (Human), and MSigDB Hallmark 2020 databases^23,24^.

To assess the cell type specificity of identified TFs and genes, we evaluated their enrichment in distinct neuronal and glial populations using cell-type markers from CellMarker 2.0^25^, a curated repository integrating data from over 100,000 publications on human and mouse tissues. This analysis enabled us to link regulatory interactions to specific brain cell types, providing additional insight into the cellular context of AD-associated transcriptional dysregulation.

### Correlation Analysis with clinical measures

To assess whether the identified regulatory edges are significantly associated with clinical measures of cognitive decline, Spearman correlation coefficients were calculated between the edge weights of the identified TF-gene interactions in the Gene Regulatory Networks (GRNs) and the estimated rates of change in various cognitive domains over time. These domains included global cognition, episodic memory, working memory, semantic memory, perceptual speed, and visuospatial ability, with the rates of change derived from regression models used in previous study^26^ that controlled for demographics or both demographics and pathology. Specifically, the following regression models were used: β Global Cognition ∼ Demographics, β Global Cognition ∼ Demographics + Pathology; β Episodic Memory ∼ Demographics, β Episodic Memory ∼ Demographics + Pathology; β Working Memory ∼ Demographics, β Working Memory ∼ Demographics + Pathology; β Semantic Memory ∼ Demographics, β Semantic Memory ∼ Demographics + Pathology; β Perceptual Speed ∼ Demographics, β Perceptual Speed ∼ Demographics + Pathology; and β Visuospatial Ability ∼ Demographics, β Visuospatial Ability ∼ Demographics + Pathology.

By assessing the strength and direction of these correlations, we aimed to find potential relationships between specific TF-gene interactions and the progression of cognitive impairment.

### Differential Expression Analysis Between AD and NCI

To identify key differentially expressed genes (DEGs) and assess their relationship to disease-specific regulatory interactions within the gene networks, we performed differential gene expression analysis between AD and NCI samples. Using pyDESeq2^27^, which implements the DESeq2 methodology within a Python framework, we normalized the expression data with Median Ratio Normalization (MRN) and modelled it using a negative binomial distribution to account for sample variability. Differential expression was assessed using a likelihood ratio test, with significance determined by an adjusted p-value (FDR) < 0.05 and a log_₂_ fold change > 0.50.

## Results

### Cohort Selection for RNA-Seq Analysis

The study included 633 participants with RNA-Seq data. Participants were categorized based on cognitive diagnosis (cogdx) as follows: 201 with no cognitive impairment (NCI), 220 with Alzheimer’s disease (AD), 33 with AD and additional cause of cognitive impairment (CI), 11 with other forms of dementia, 158 with mild cognitive impairment (MCI) without another cause of CI, and 10 with MCI and another cause of CI. Neuropathological classification (CERAD) further categorized participants as 184 definite AD, 218 probable AD, 67 possible AD, and 164 with no AD. To ensure diagnostic specificity and minimize confounds, we retained only cases meeting CERAD criteria for Definite AD and cogdx classification of AD (no alternative cause of CI) as AD cases. NCI controls were selected based on the absence of neuropathological (CERAD) and clinical (cogdx) evidence of AD. This resulted in a final cohort of 110 AD cases and 78 NCI controls for downstream analysis.

### Preprocessed RNA-Seq expression for Gene Regulatory Network Construction

After applying the CPM threshold filter to the 110 AD and 78 NCI samples, the dataset was refined to include 17,330 genes that met the expression criteria, retaining genes that are consistently highly expressed across both the AD and NCI groups. To address potential batch effects, we employed Combat-Seq, a technique specifically designed for RNA-Seq data. Following this correction, we observed that batches 0 and 7 still had a significant impact on the expression counts. As a result, we excluded batches 0 and 7 from the analyses due to their persistent effects on gene expression despite the batch correction steps. Furthermore, we applied quantile-based thresholds (5th and 95th percentiles) to filter out extreme samples based on their principal component analysis (PCA) components. By the end of preprocessing pipeline, we retained 87 AD samples and 67 NCI samples for downstream analysis.

### Demographic and Clinical Characteristics

Participants with AD (n = 87) were marginally older than those with NCI (n = 67), with mean ages of 89.8 years (SD = 5.6) and 84.3 years (SD = 7.2), respectively, and an independent t-test revealed a significant age difference (p-value = 1.11e-06). Among AD cases, 56 were female and 31 were male, while in the NCI group, 36 were female and 31 were male. A Chi-square test indicated no significant difference in gender distribution between the two groups (χ^2^ = 1.37, p = 0.24). APOE genotype distribution in the NCI group was as follows: ε2/ε2 (n=1), ε2/ε3 (n=15), ε2/ε4 (n=1), ε3/ε3 (n=47), and ε3/ε4 (n=3). In the AD group, the APOE genotype distribution was ε2/ε2 (n=4), ε2/ε3 (n=3), ε2/ε4 (n=42), ε3/ε3 (n=37), and ε3/ε4 (n=1). A Chi-square test comparing APOE genotypes between the AD and NCI groups revealed a significant difference in APOE genotype distribution (p = 5.01e-10).

The global AD pathology burden (gpath), a quantitative measure summarizing AD pathology, was significantly higher (t-test, p-value=8.12e-38) in AD cases (mean = 1.48, SD = 0.59) compared to NCI controls (mean = 0.11, SD = 0.14). Gpath is derived from counts of three AD pathologies, neuritic plaques (n), diffuse plaques (d), and neurofibrillary tangles (nft), assessed through microscopic examination of silver-stained slides from five brain regions: midfrontal cortex, midtemporal cortex, inferior parietal cortex, entorhinal cortex, and hippocampus (CA1)^28^.

### Network Metrics Differences in AD-Specific and NCI-Specific GRNs

To gain a systems-level understanding of AD, we constructed sample-specific GRNs for each of the 87 AD and 67 NCI samples, leveraging multi-omics data from postmortem brain tissue. This individualized network approach captured inter-individual variability in transcriptional regulation, potentially reflecting differences in molecular pathology. Our analysis, using the non-parametric Mann-Whitney U test, did not reveal a significant difference in density between AD-specific and NCI-specific GRNs (p-value = 0.19). Similarly, no significant differences were observed in the average degree (p-value = 0.17) or modularity (p-value = 0.10). However, we identified a notable difference in the largest connected component size (p-value = 0.025), with AD showing a noticeably smaller largest connected component than NCI, indicating a structural variation in network connectivity. This variation may reflect changes in central regulatory modules and thus key biological processes within the network. Additionally, nestedness showed a significant difference (p-value < 0.05), with NCI GRNs has higher nestedness than AD, highlighting variations in the connectivity patterns between nodes of different degrees. These findings highlight key network properties that may be altered in the context of AD. This information is summarized in Table 1. (Box plots of network metrics in AD and NCI are shown in supplementary Figure S2). These findings highlight key network properties that may be altered in the context of AD.

**Table 1.** AD and NCI GRNs structural features.

| Metric | AD (Mean ± Std) | NCI (Mean ± Std) | P-value |
| --- | --- | --- | --- |
| Density | 0.51 ± 0.01 | 0.51 ± 0.008 | 0.19 |
| Average Weight Degree | 802.67 ± 332.4 | 805.91 ± 239.5 | 0.17 |
| Modularity | 0.21 ± 0.03 | 0.22 ± 0.03 | 0.1 |
| Giant Component Size | 17188.10 ± 23.51 | 17192.19 ± 23.03 | 0.025 |
| Nestedness | 0.52 ± 0.04 | -0.50 ± 0.04 | 0.046 |

### Sample-Specific Gene Regulatory Networks Reveal Key Regulatory Differences in AD

To identify key regulatory differences, we applied logistic regression with Recursive Feature Elimination (RFE), achieving a weighted classification accuracy of 96% based on TF-gene edge weights. RFE identified 22 critical TF-gene interactions that effectively distinguish AD from NCI, with cross-validation confirming model robustness (weighted accuracy: 96 ± 0.2%, F1-score: 0.95 ± 0.5). These key interactions provide insights into AD’s regulatory mechanisms and potential biomarkers for diagnosis. A confusion matrix [Figure 2A] illustrates the classification performance on cross validation test set, while t-SNE plots [Figure 2B] of the 22 key TF-target gene interactions show clear separation between AD and NCI samples. Bootstrapping analysis, in which random TF-target gene pairs were selected, demonstrated that the 96% weighted accuracy observed using the 22 key interactions was statistically significant (p-value < 0.05), indicating that this high classification performance was unlikely due to chance. These results underscore the predictive power of transcriptional regulation in differentiating AD from NCI, suggesting a clear disease-related network disruption and biological relevance of the identified TF-gene interactions in AD pathology.

**Figure 2.**
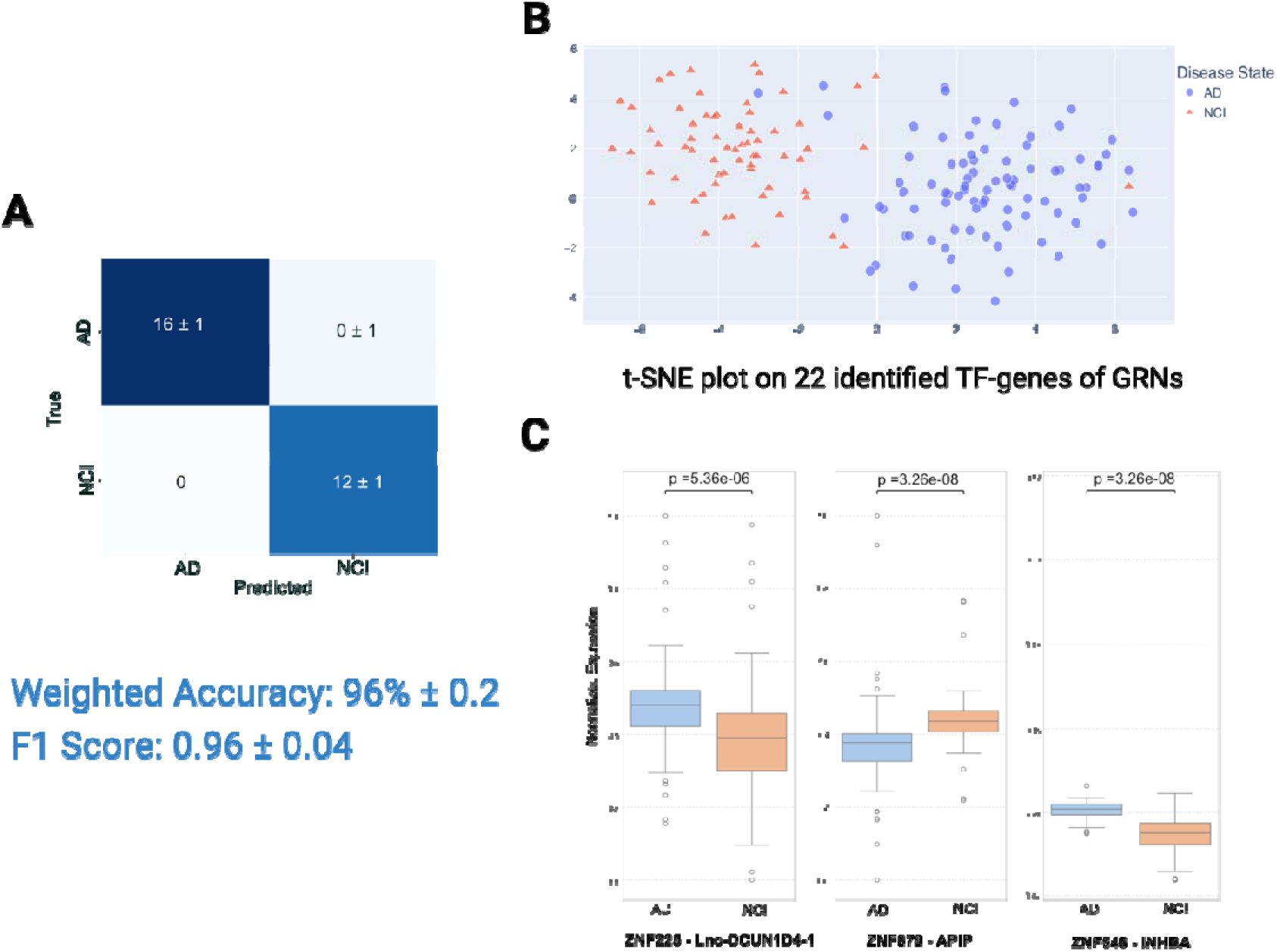
Results of Logistic Regression for classifying Alzheimer’s Disease (AD) and Mild Cognitive Impairment (NCI) Gene Regulatory Networks (GRNs) A. Confusion matrix illustrating the performance of Logistic Regression in classifying Alzheimer’s Disease (AD) and Mild Cognitive Impairment (NCI) GRNs. B. t-SNE plot showing the separation of Alzheimer’s Disease (AD) and Mild Cognitive Impairment (NCI) Gene Regulatory Networks (GRNs) based on 22 identified GRN edges from Logistic Regression with Recursive Feature Elimination (RFE) C. Box plot illustrating the distribution of three hub TF-gene edge weights present in the 22 identified TF-gene edges for the Mild Cognitive Impairment (NCI) and Alzheimer’s Disease (AD) groups.

To determine whether any of the 22 identified TF–gene interactions represented highly influential regulators, we assessed which transcription factors functioned as regulatory hubs. A transcription factor was defined as a hub if its connectivity, measured as the number of downstream targets, was at least twice the average degree of all transcription factors in the network. This criterion highlighted TFs with disproportionate regulatory influence within the network. Based on this criterion, we identified three transcription factors, namely ZNF225, ZNF849, and ZNF548, as hub nodes within the GRNs among the 22 key TF-gene interactions. These hubs form key regulatory pairs with Lnc-DCUN1D4-1, APIP, and INHBA, respectively, within the identified 22 TF-gene interactions. The differences in the weights of these three TF-gene regulatory interactions between AD and NCI are illustrated in Figure 2C.

### Identification of Dysregulated Pathways in AD through Gene Set Enrichment Analysis

To investigate biological processes associated with AD, we performed Gene Set Enrichment Analysis (GSEA) on differentially expressed genes from AD and NCI samples. This analysis identified 88 significantly enriched pathways (FDR-adjusted p-value < 0.005) [Figure 3] spanning multiple biological functions relevant to neurodegeneration and AD pathophysiology. A comprehensive list of enriched pathways identified is provided in Supplementary Table S1.

**Figure 3.**
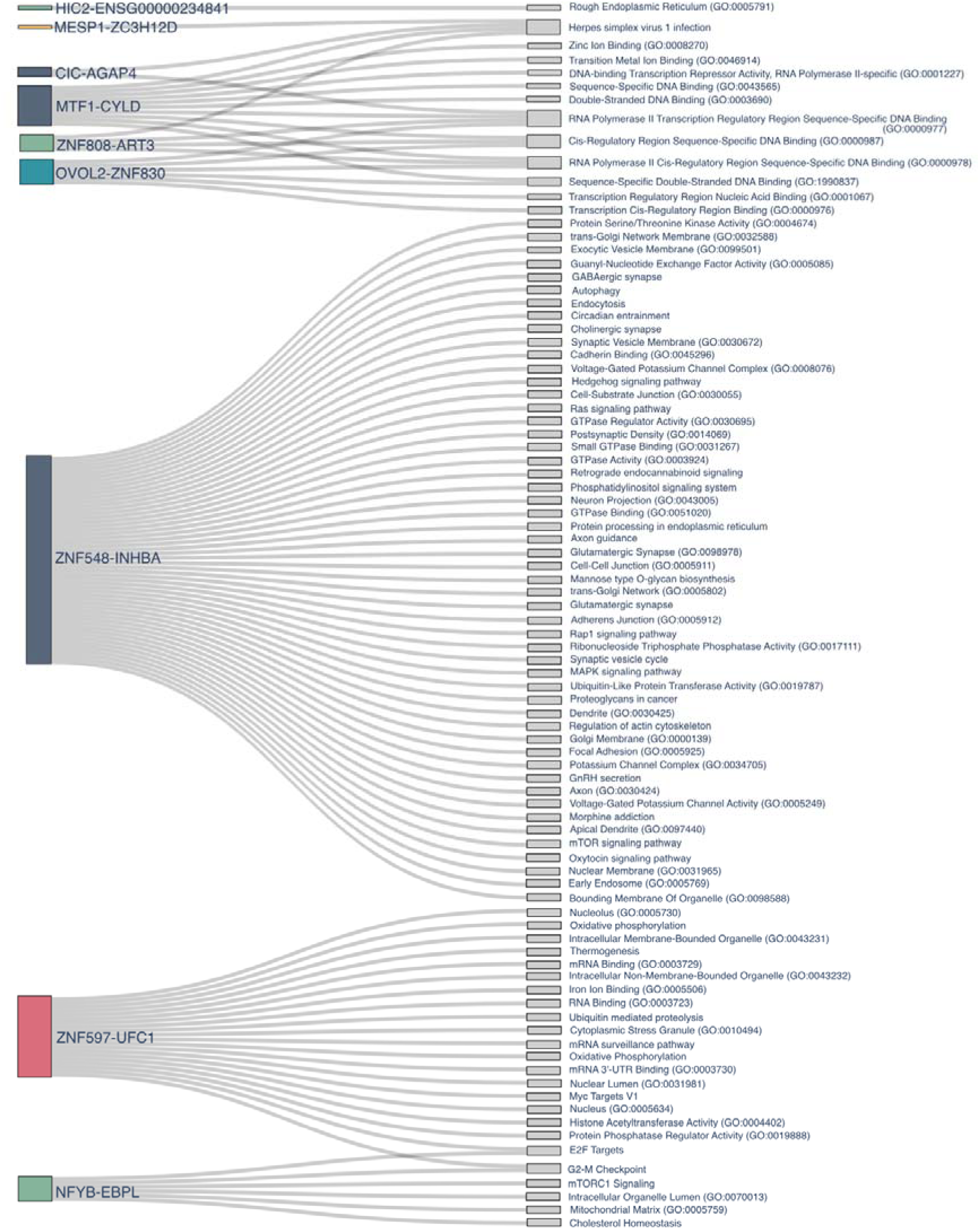
Sankey Plot of TF-Gene Regulatory Interactions and Enriched Pathways. Sankey plot illustrating transcription factor (TF)-gene regulatory edges and their associated genes enriched in biological pathways. The plot visualizes the flow of regulatory interactions, highlighting significantly enriched pathways with a false discovery rate (FDR) p-value < 0.005.

Pathways associated with synaptic function and neurotransmission, including cholinergic synapse, glutamatergic synapse, GABAergic synapse, synaptic vesicle cycle, and postsynaptic density, were significantly enriched in AD samples. Disruptions in protein homeostasis and autophagy were also prominent, with ubiquitin-mediated proteolysis, protein processing in the endoplasmic reticulum, mTOR signaling, and autophagy showing enrichment.

GSEA further identified dysregulation in oxidative stress and mitochondrial function-related pathways, including oxidative phosphorylation, mitochondrial matrix components, and transition metal ion binding. Mitochondrial dysfunction and oxidative stress contribute to neuronal damage and energy deficits in AD. Additionally, neuroinflammatory processes and cell adhesion mechanisms were enriched, with pathways such as regulation of actin cytoskeleton, focal adhesion, cell-cell junctions, MAPK signalling, and Ras signalling showing altered activity.

In terms of molecular functions, our analysis identified enrichment in GTPase activity, GTPase binding, guanyl-nucleotide exchange factor activity, protein serine/threonine kinase activity, ubiquitin-protein transferase activity, voltage-gated potassium channel activity, RNA binding, and iron ion binding.

The analysis also revealed enrichment in cellular components, including Golgi membrane, trans-Golgi network, mitochondrial matrix, rough endoplasmic reticulum, nucleus, postsynaptic density, synaptic membrane, neuron projection, and axon guidance.

Additionally, key transcriptional and gene regulatory pathways were identified, including E2F targets, Myc targets V1, histone acetyltransferase activity. sequence-specific DNA binding, RNA polymerase II transcription regulatory region sequence-specific DNA binding, and transcription cis-regulatory region binding.

### Cell-Type Associations Highlight the Role of Immune Cells

We conducted a cell-type association analysis to investigate the contribution of specific cell populations to AD. We identified ATOH8 as a marker of microglia, NEUROD1 as a marker of neurons, and TCF7 in macrophages, overlapping with the 22 TF-gene edges. This finding shows the involvement of immune response (microglia and macrophages) and neuronal function, with key 22 TF-gene edges in Alzheimer’s Disease.

### Key Regulatory Interactions with Previously Studied Alzheimer’s Disease Genes

To determine whether our identified 22 key TF-gene regulatory interactions influence or regulate other AD-related genes, we examined their association with previously studied AD-targeted genes curated by researchers from the National Institute on Aging’s Accelerating Medicines Partnership in Alzheimer’s Disease (AMP-AD) consortium30. We systematically assessed whether these genes were regulated by or interacted with the identified TF-gene interactions in the GRNs. Notably, over half of the AD-targeted genes were regulated by or interacted with these 22 regulatory edges [Figure 4], suggesting a high relevance of the 22 TF-genes in AD pathophysiology and in the molecular mechanisms underlying the disease.

**Figure 4.**
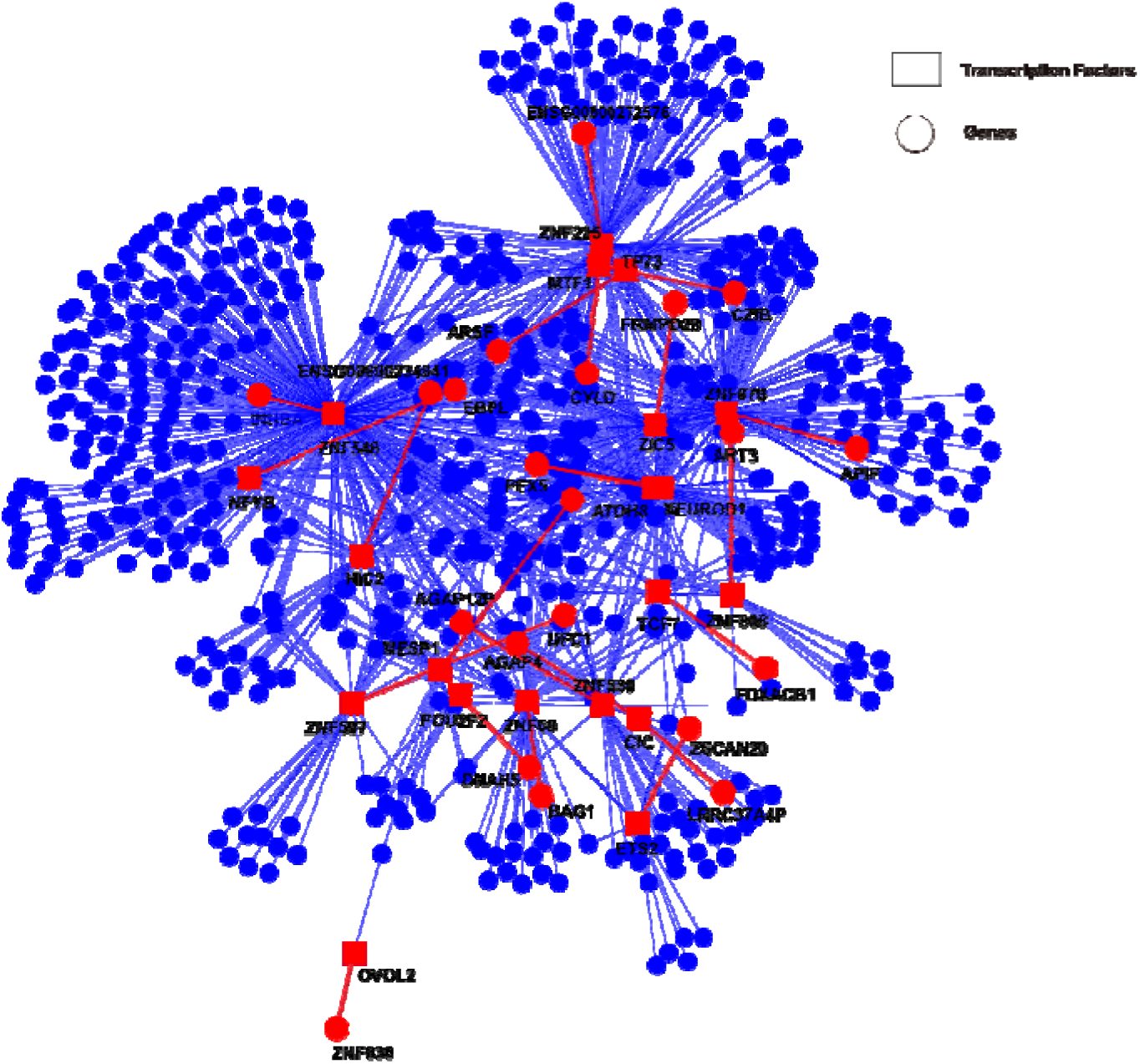
Network plot illustrating the 22 identified TF-gene regulatory edges and their associations with AD-targeted genes from the AMP-AD consortium. The plot highlights the significant interactions and regulatory relationships, underscoring the importance of these edges in Alzheimer’s disease pathophysiology

### Differential Expression Analysis

Differential expression analysis identified a total of 74 significant DEGs between the AD and NCI samples, using a threshold of adjusted p-value (FDR) < 0.05 and absolute log_₂_ fold change > 0.50 [supplementary Figure S4.A] Among these, only one gene, FRMPD2B, overlapped with significant transcription factor (TF)-gene edges in the regulatory network [supplementary Figure S4.B] This finding highlights the distinct insights gained from gene regulatory network (GRN) analysis compared to traditional differential expression analysis. To further investigate the functional relevance of these DEGs, we performed Gene Set Enrichment Analysis (GSEA) using multiple curated gene sets: GO Molecular Function 2023, MSigDB Hallmark 2020, KEGG 2021 (Human), GO Cellular Component 2023. The analysis identified enrichment in key biological processes, including TNF-alpha signaling via NF-kB, structural components like collagen-containing extracellular matrix (GO:0062023) and microvillus (GO:0005902), as well as molecular activities such as hormone activity (GO:0005179) and neutral L-amino acid:sodium symporter activity (GO:0005295).

## Discussion

This study aimed to explore transcriptional network dysregulation in Alzheimer’s Disease (AD) through the analysis of individual-specific gene regulatory network models. While previous studies have extensively cataloged individual risk genes and patterns of gene-expression changes, the role of disruptions of the interactions among transcription factors (TFs) and their regulatory targets in AD remained unknown. By employing a systems biology approach to model individual-specific genome-scale gene regulatory networks, we addressed this critical gap, uncovering AD-specific regulatory dynamics and identifying transcription factor-gene interactions significantly associated with declines in cognition and memory.

In our analysis, we found significant differences in the regulatory networks between individuals diagnosed with AD and those without cognitive impairment (NCI). Specifically, 22 critical TF-gene interactions reliably distinguished AD from NCI with a classification accuracy of 96%. Among these interactions, ZNF225, ZNF849, and ZNF548 emerged as key hub transcription factors, exerting substantial influence across the networks. Thus, specific disruptions in TF-gene regulatory networks appear to have meaningful clinical relevance, suggesting a potential relevance as biomarkers or therapeutic targets.

Our findings support the hypothesis that AD is a highly complex disease characterized by multiple interconnected regulatory interactions collectively contributing to its molecular pathology. The prominence of zinc-finger transcription factors as regulatory hubs aligns with existing literature implicating these proteins in neural differentiation, synaptic plasticity, and neurodegeneration^29^. Our network-based approach expands upon the insights obtained from traditional differential expression analyses by adding crucial functional information, identifying specific transcription factors within patient-specific gene regulatory networks that are directly associated with cognitive decline in Alzheimer’s disease.

Moreover, our gene set enrichment analysis identified significant dysregulation in key biological pathways critical to neuronal function in AD, including synaptic transmission, mitochondrial energy metabolism, protein homeostasis, neuroinflammation, and epigenetic regulation. Synaptic dysfunction was particularly prominent, with disturbances in cholinergic, glutamatergic, and GABAergic pathways, all of which are closely linked to synapse loss and cognitive decline in AD^30–32^. In parallel, our analysis revealed marked mitochondrial dysfunction and oxidative stress, with disruptions in energy metabolism and oxidative phosphorylation leading to decreased ATP production, elevated reactive oxygen species (ROS), and neuronal damage. These findings align with the mitochondrial cascade hypothesis^33^, wherein impaired mitochondrial function amplifies amyloid-β (Aβ) and tau pathology, creating a feedback loop of oxidative damage and synaptic failure that accelerates AD progression. Protein homeostasis failure also emerged as dysregulated in our analysis, with alterations in critical pathways responsible for protein quality control, including the ubiquitin proteasome system (UPS), endoplasmic reticulum (ER) protein processing, autophagy, and mTOR signaling. These disruptions are assumed to overwhelm normal protein clearance mechanisms, leading to the accumulation of toxic Aβ plaques and tau tangles^34^. The impairment of proteasomal and autophagic pathways, coupled with hyperactivation of mTOR signaling, contributes to the buildup of amyloid and tau, core drivers of AD pathology^35–37^. Neuroinflammation was another key feature identified in our analysis, with significant upregulation of cytokine signaling, complement activation, and glial markers, reflecting chronic activation of microglia and astrocytes. This persistent neuroinflammation, combined with blood-brain barrier dysfunction, worsens neurodegeneration, as thought to impair Aβ clearance, and accelerate AD progression^38–40^. Taken together, these findings are consistent with the concept of “inflammaging,” where chronic, low-grade inflammation in the brain exacerbates neurodegeneration and contributes to AD pathology^40^. Finally, significant transcriptional and epigenetic dysregulation was observed in AD, with alterations in histone acetylation, chromatin organization, and transcription factor binding, particularly involving key regulators such as E2F and Myc. These changes suggest aberrant reactivation of the cell cycle and stress response pathways in neurons, potentially contributing to neurodegeneration. Epigenetic modifications, including chromatin remodeling and disruptions in DNA methylation, exacerbate disease pathology by altering gene expression, which may silence protective genes and activate harmful pathways^41^. Together, these transcriptional regulation shifts highlight the synergistic interplay of multiple mechanisms in AD, including synaptic failure, mitochondrial dysfunction, proteostasis collapse, neuroinflammation, and epigenetic alterations, likely all acting together during the progression of the disease.

Several limitations warrant consideration. Our moderate sample size may limit generalizability, and the reliance on postmortem data introduces inherent variability despite rigorous normalization. The cross-sectional nature of post-mortem tissue precludes direct assessments of causality or longitudinal changes in gene regulatory dynamics. Future research is needed to address these limitations by validating identified regulatory interactions in larger independent cohorts and employing longitudinal designs coupled with serial cognitive assessments.

Further mechanistic validation, including experimental modulation of identified hub genes through gene-editing technologies and animal models, will be essential to delineate their precise roles in AD pathology. Moreover, expanding analyses beyond the dorsolateral prefrontal cortex to other vulnerable brain regions such as the hippocampus and entorhinal cortex could provide additional insights. Single-cell and spatial transcriptomic approaches may further refine our understanding of cell-type-specific regulatory changes in AD.

Despite these limitations, the strength of our study lies in keeping individual differences in our analysis by reconstructing personalized Gene Regulatory Networks (GRNs). This approach allows for a deeper understanding of gene regulatory interactions that may be missed in group-based analyses, providing more specific insights into the molecular mechanisms of AD. Additionally, by using GRNs, which model TF-gene interactions rather than relying solely on gene expression values, we focus on the regulatory relationships that drive disease processes, offering a more functional perspective on AD.

In conclusion, this study identifies key transcriptional hubs within individual-level gene regulatory networks significantly associated with Alzheimer’s disease. By highlighting network dysregulation rather than isolated genetic alterations, our findings offer novel insights into the molecular complexity of AD and open promising avenues for personalized therapeutic interventions targeting regulatory hubs such as ZNF225, ZNF849, and ZNF548. Ultimately, these insights advance our understanding of AD’s molecular underpinnings and provide a framework for future targeted therapeutic strategies.

### Code availability

Python codes for all analysis are available on a GitHub public repository: https://github.com/Polster-lab/Analysis_GRN_ROSMAP

### Author contributions

DA: Data analysis, Writing, Interpretation; EK: Interpretation, Writing; AP: Conceptualization, Data curation, Formal analysis, Investigation, Methodology, Software, Validation, Visualization, Writing

## Data Availability

All data produced are available online at synapse.org

https://www.synapse.org/Synapse:syn3219045

## Acknowledgements

The results published here are in whole or in part based on data obtained from the AD Knowledge Portal (https://adknowledgeportal.org). Study data were provided by the Rush Alzheimer’s Disease Center, Rush University Medical Center, Chicago. Data collection was supported through funding by NIA grants P30AG10161 (ROS), R01AG15819 (ROSMAP; genomics and RNAseq), the Illinois Department of Public Health (ROSMAP), and the Translational Genomics Research Institute (genomic). Additional phenotypic data can be requested at www.radc.rush.edu. The data handling was enabled by resources provided by the National Academic Infrastructure for Supercomputing in Sweden (NAISS), partially funded by the Swedish Research Council through grant agreement no. 2022-06725.

## Supplementary figures

**Figure S1.**
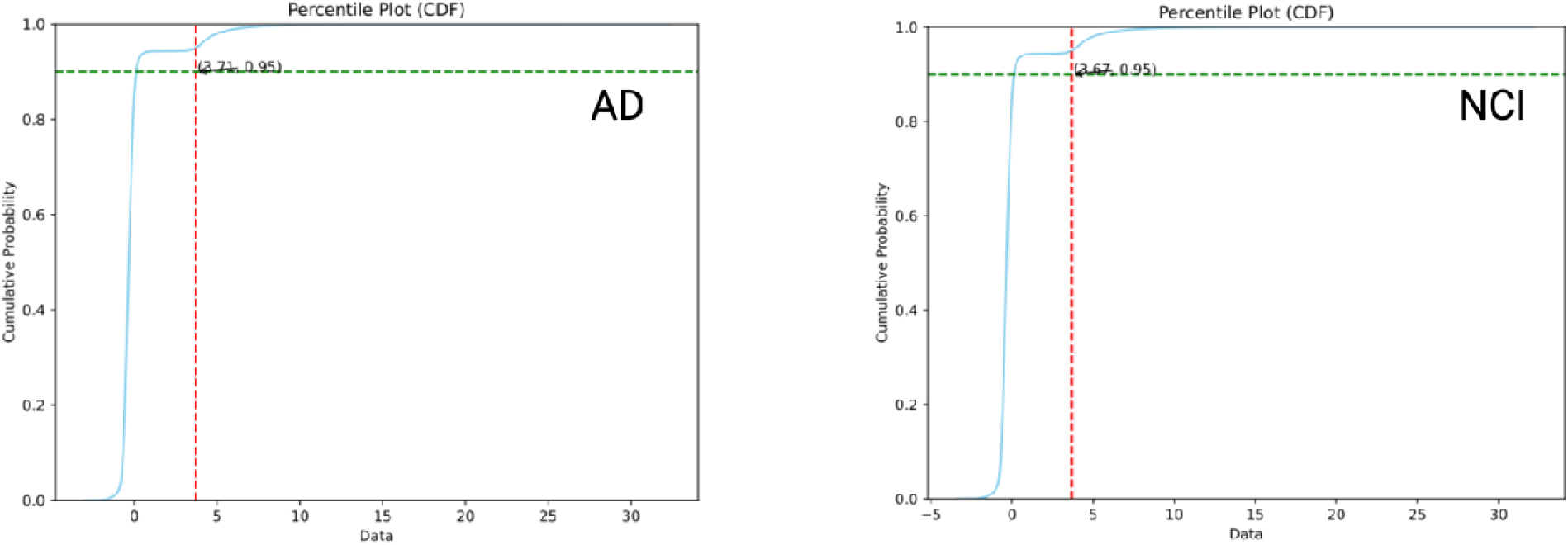
Cumulative distribution of edge weights for Alzheimer’s Disease (AD) and Mild Cognitive Impairment (NCI) networks, highlighting the threshold selection for the top 5% of regulatory interactions.

**Figure S2.**
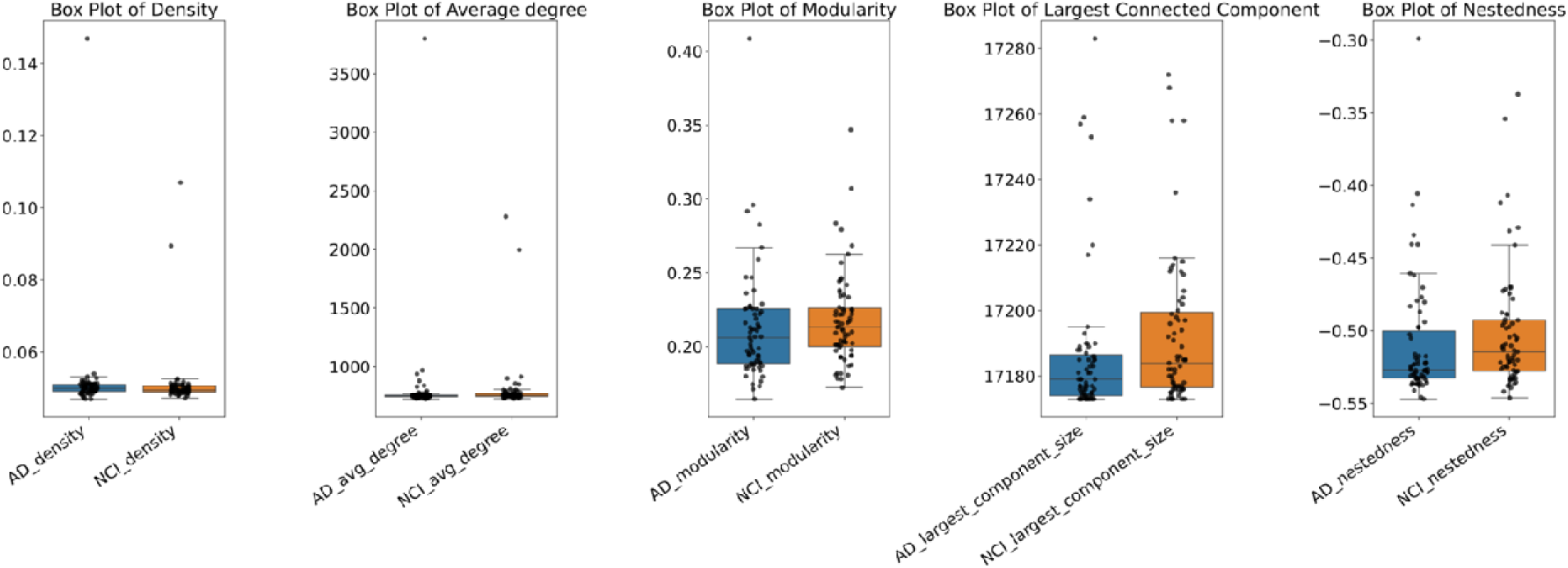
Box plots showing network metrics (density, average degree, modularity, giant component size, and nestedness) in Alzheimer’s Disease (AD) and Mild Cognitive Impairment (NCI) networks.

**Figure S3.**
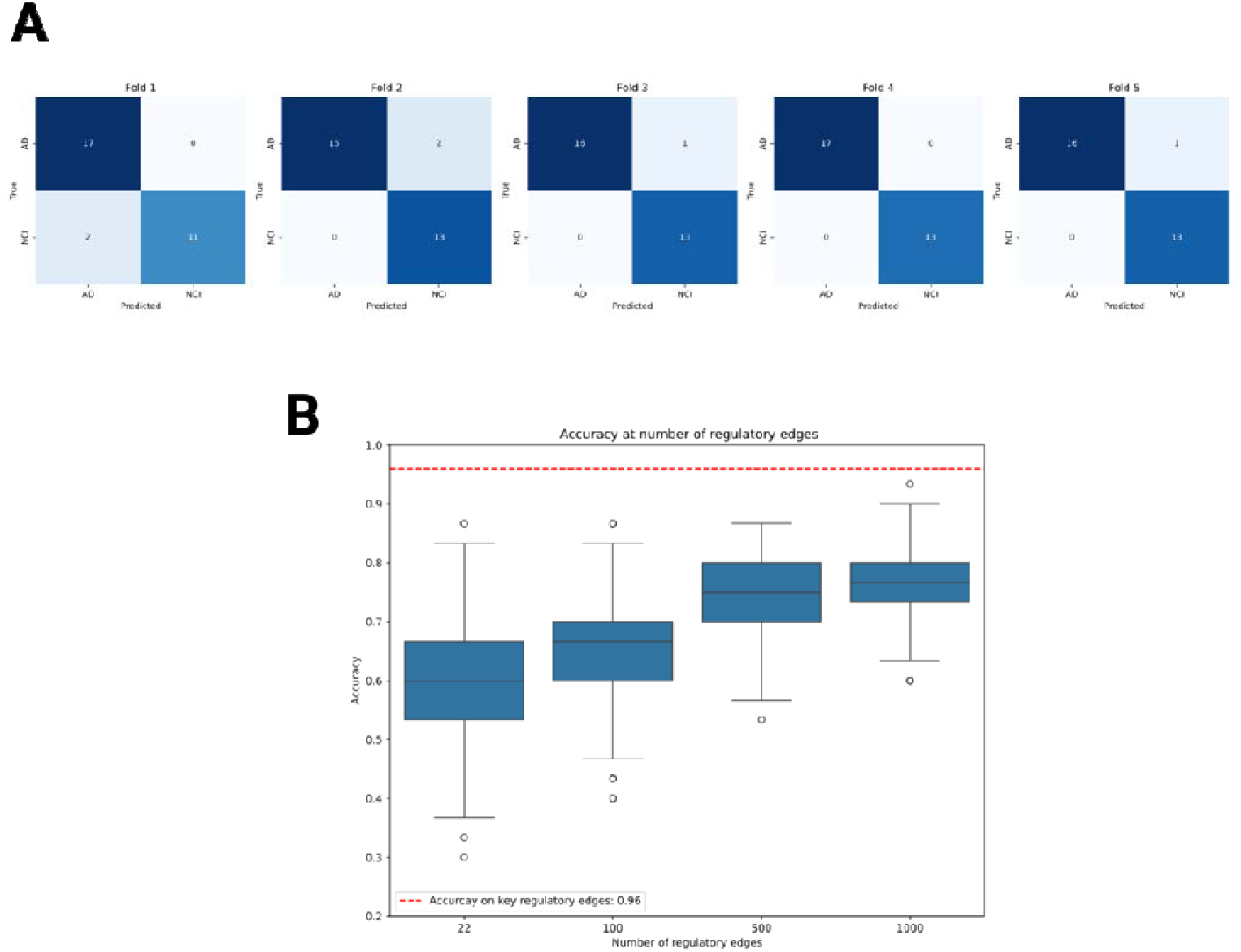
A. 5-fold cross-validation confusion matrix showing the performance of classifying Alzheimer’s Disease (AD) and No Cognitive Impairment (NCI) using 22 identified TF-gene edges B. Box plots showing the classification accuracy at different levels of random TF-target gene pair selection (22, 100, 500, and 1000 gene pairs) across 1000 bootstrapping iterations.

**Figure S4.**
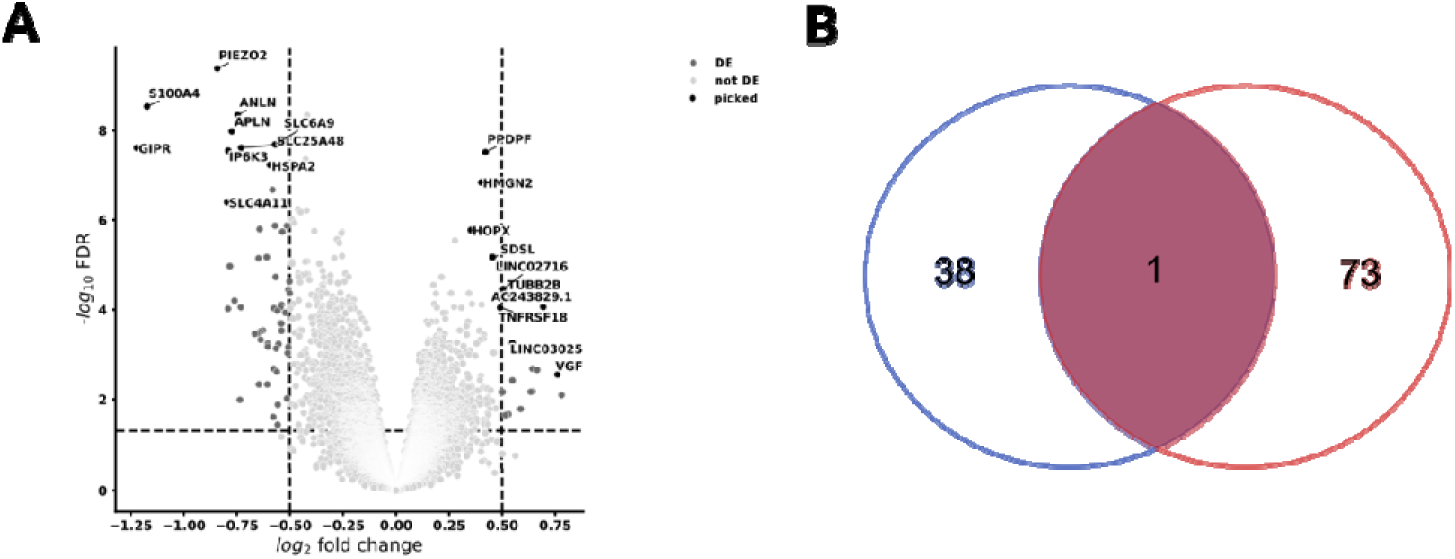
A. Volcano plot of differential expression analysis highlighting significantly differentially expressed genes (DEGs). B. Venn diagram showing the overlap between DEGs and the 22 identified TF-gene edges, illustrating the intersection of key regulatory interactions with differentially expressed genes.

## Supplementary Tables

**Table S1.**
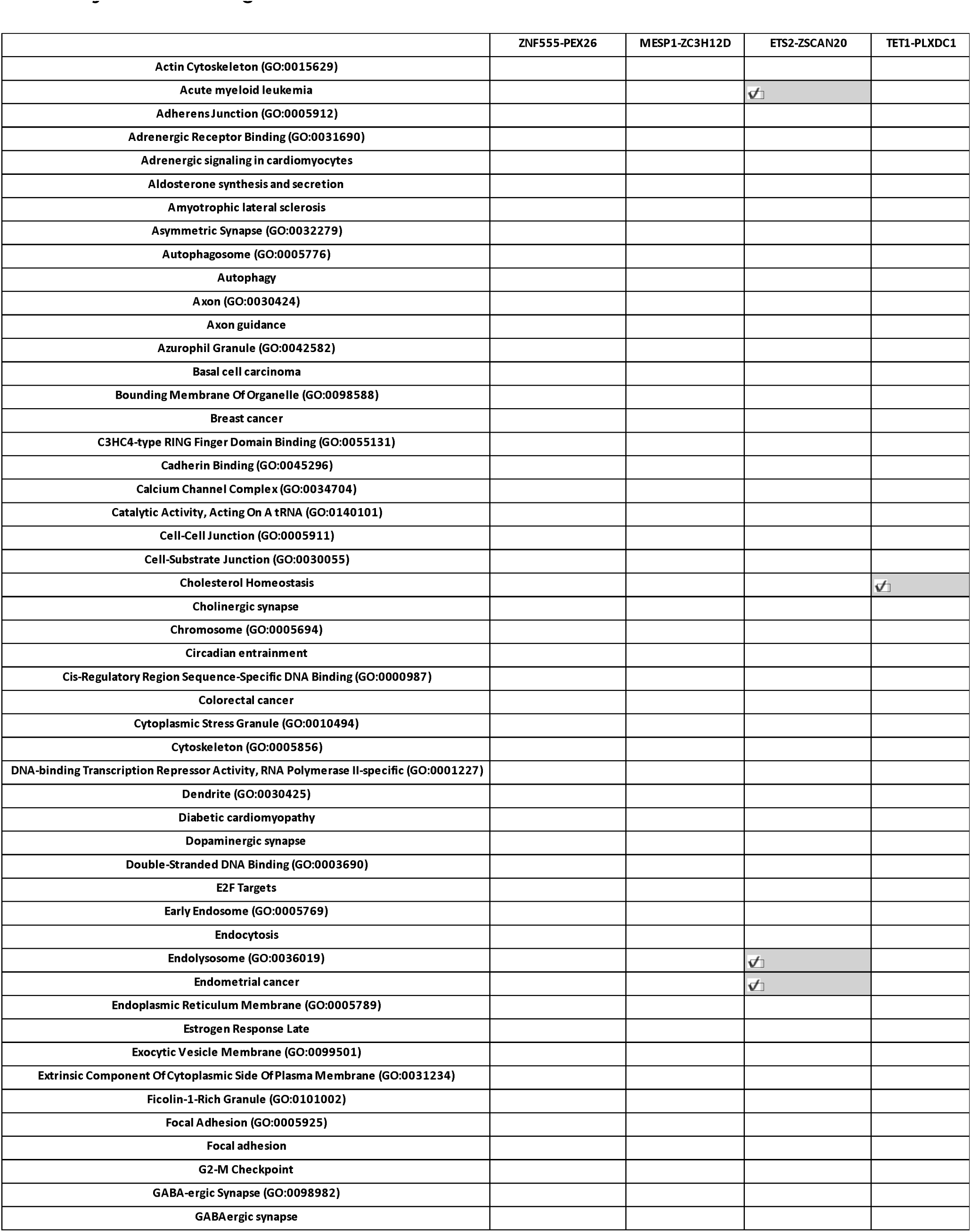
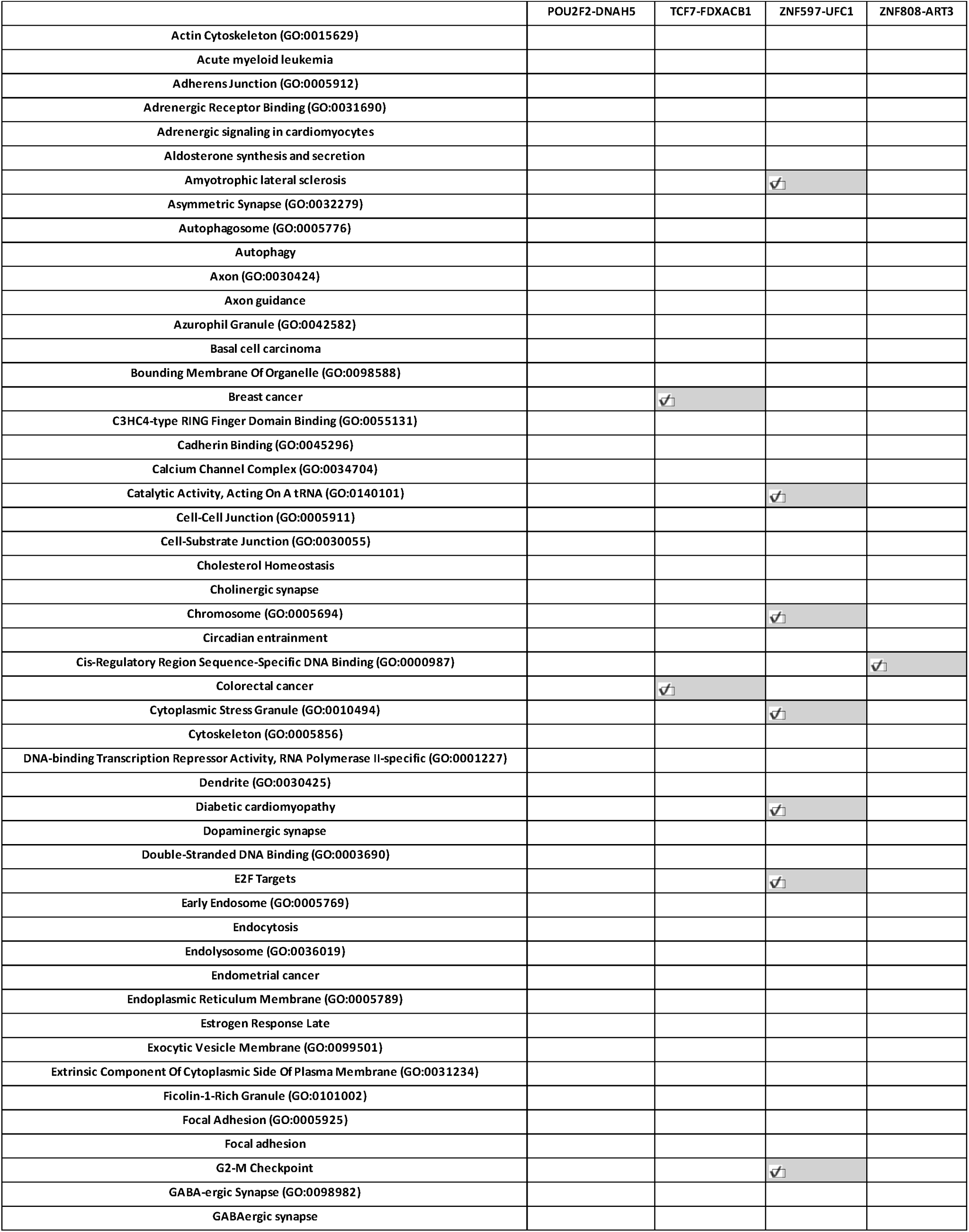
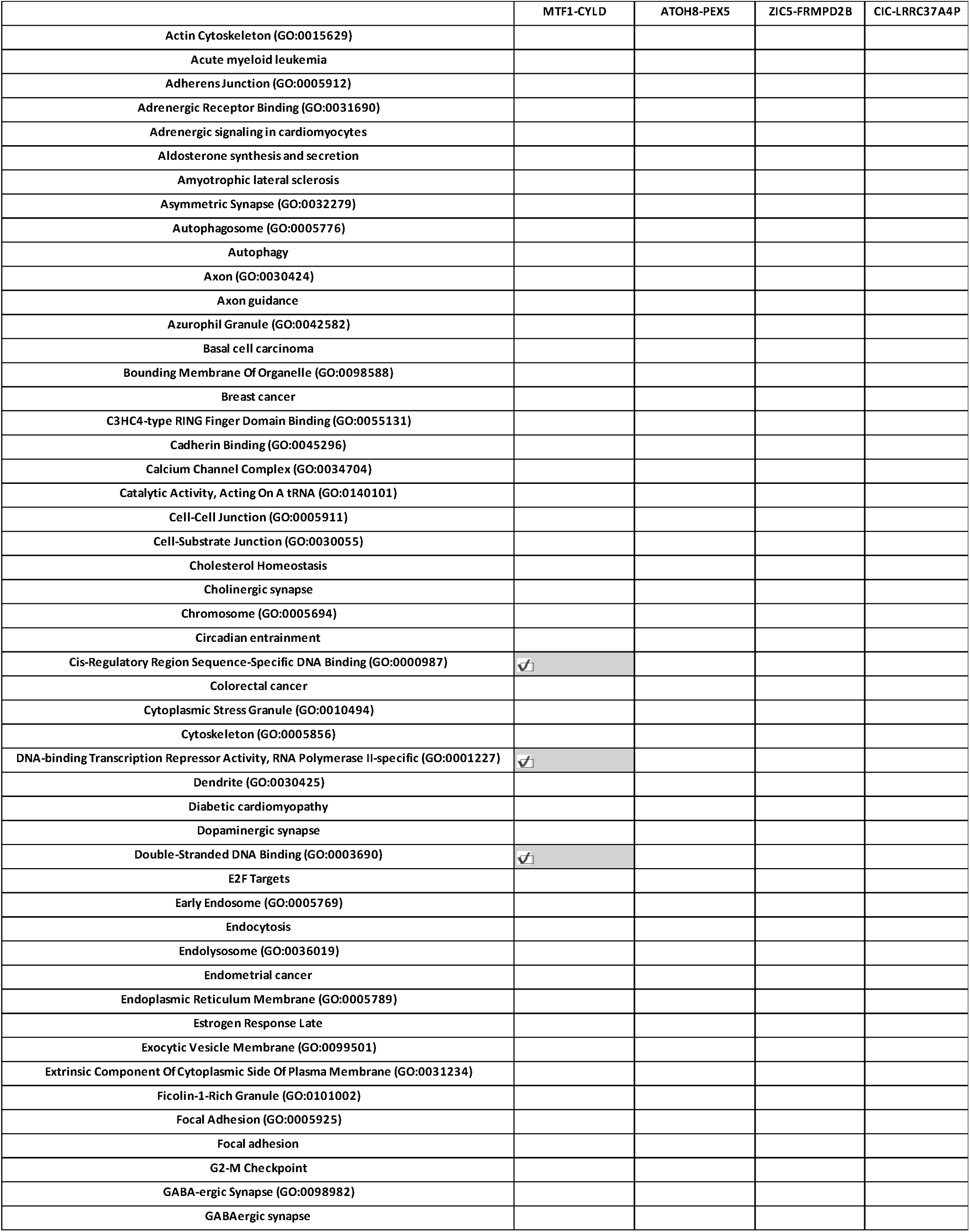
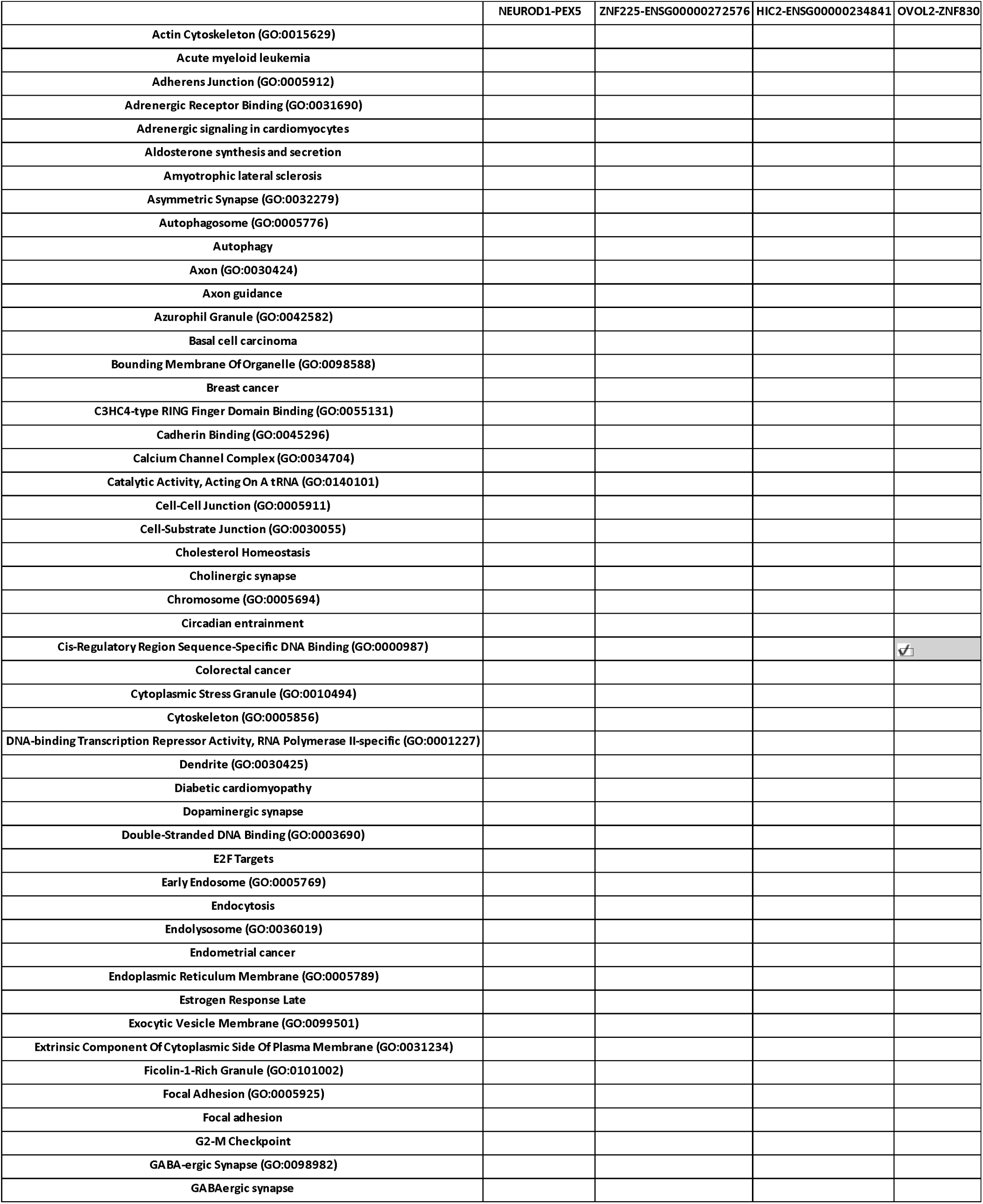
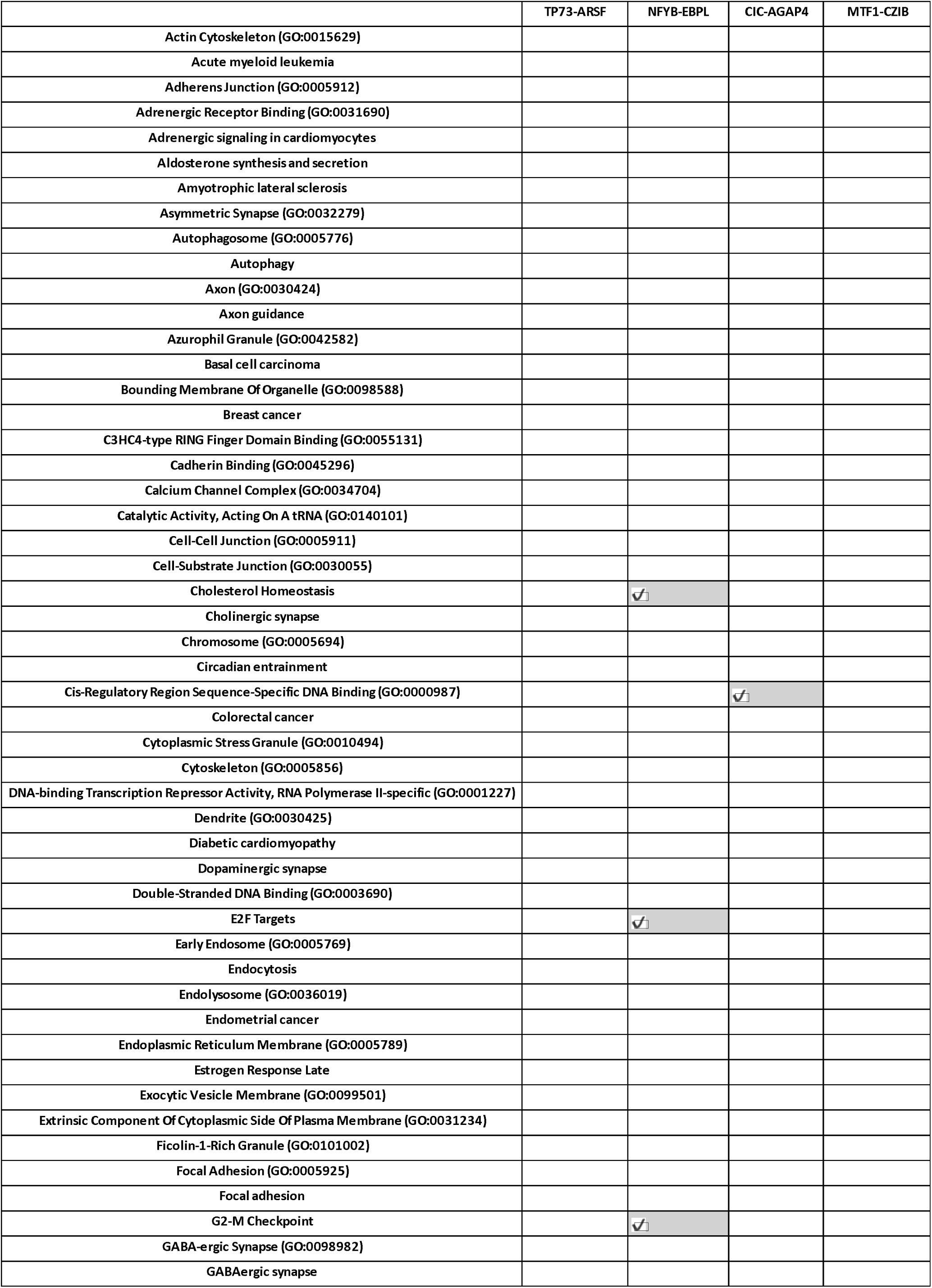
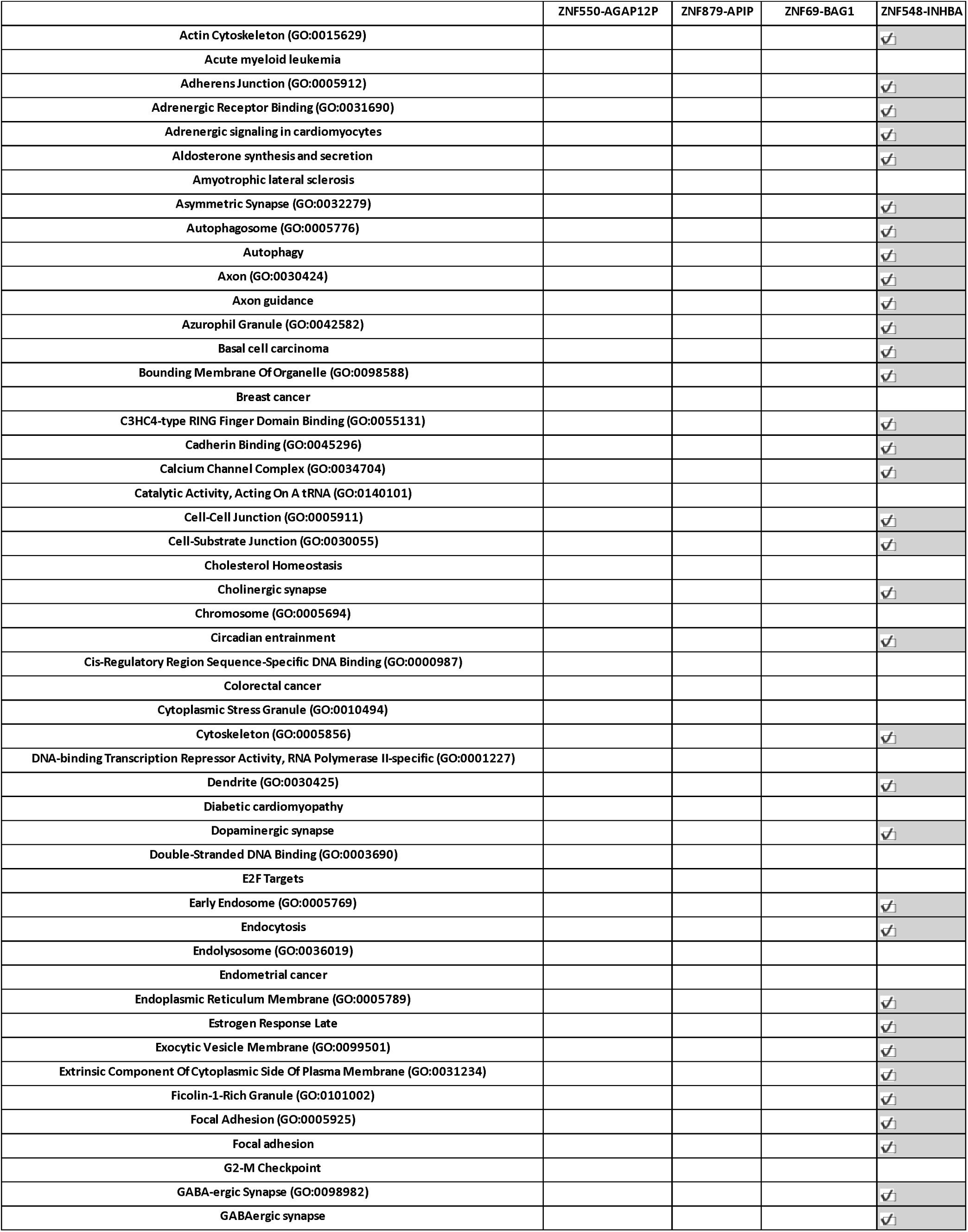
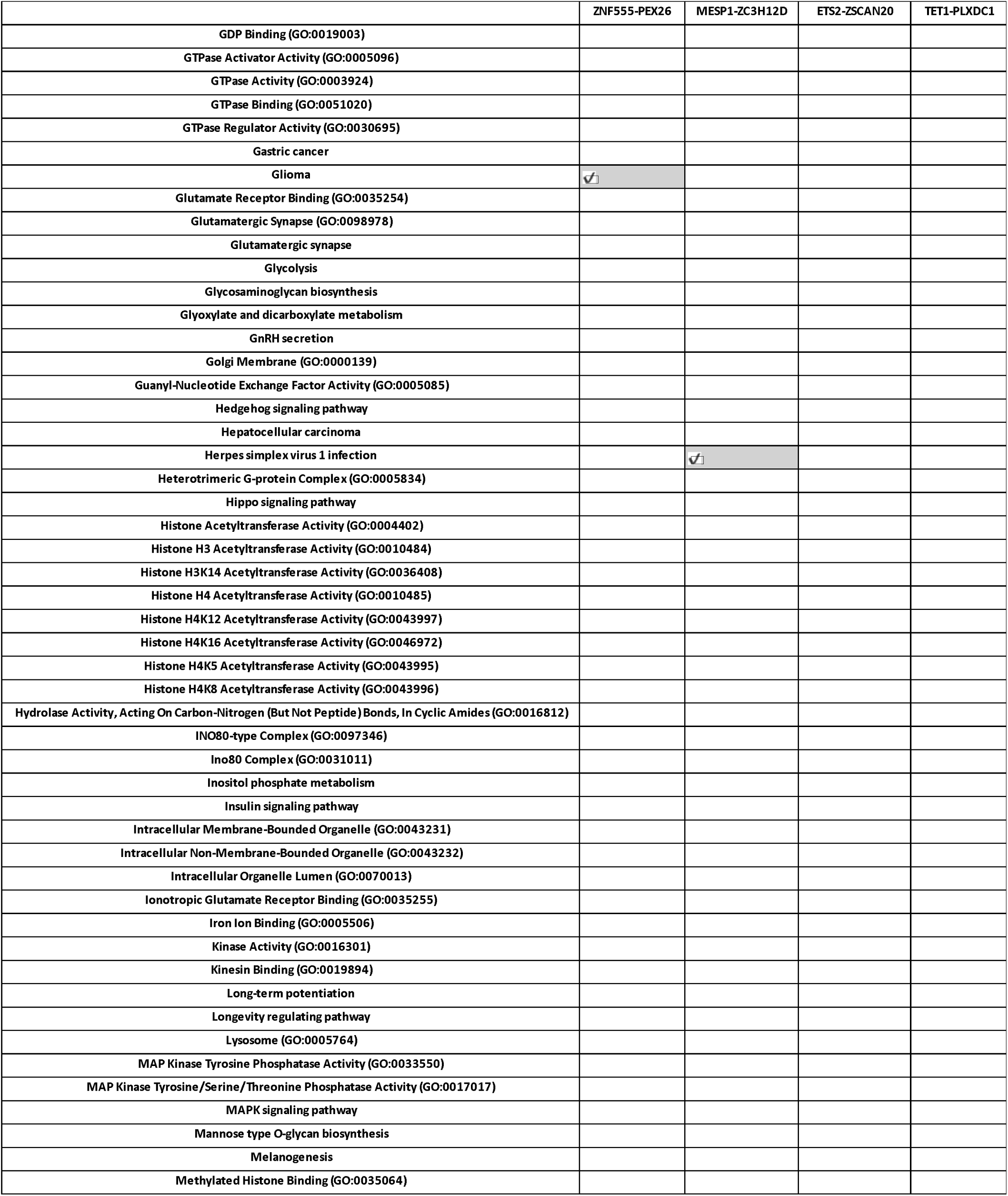
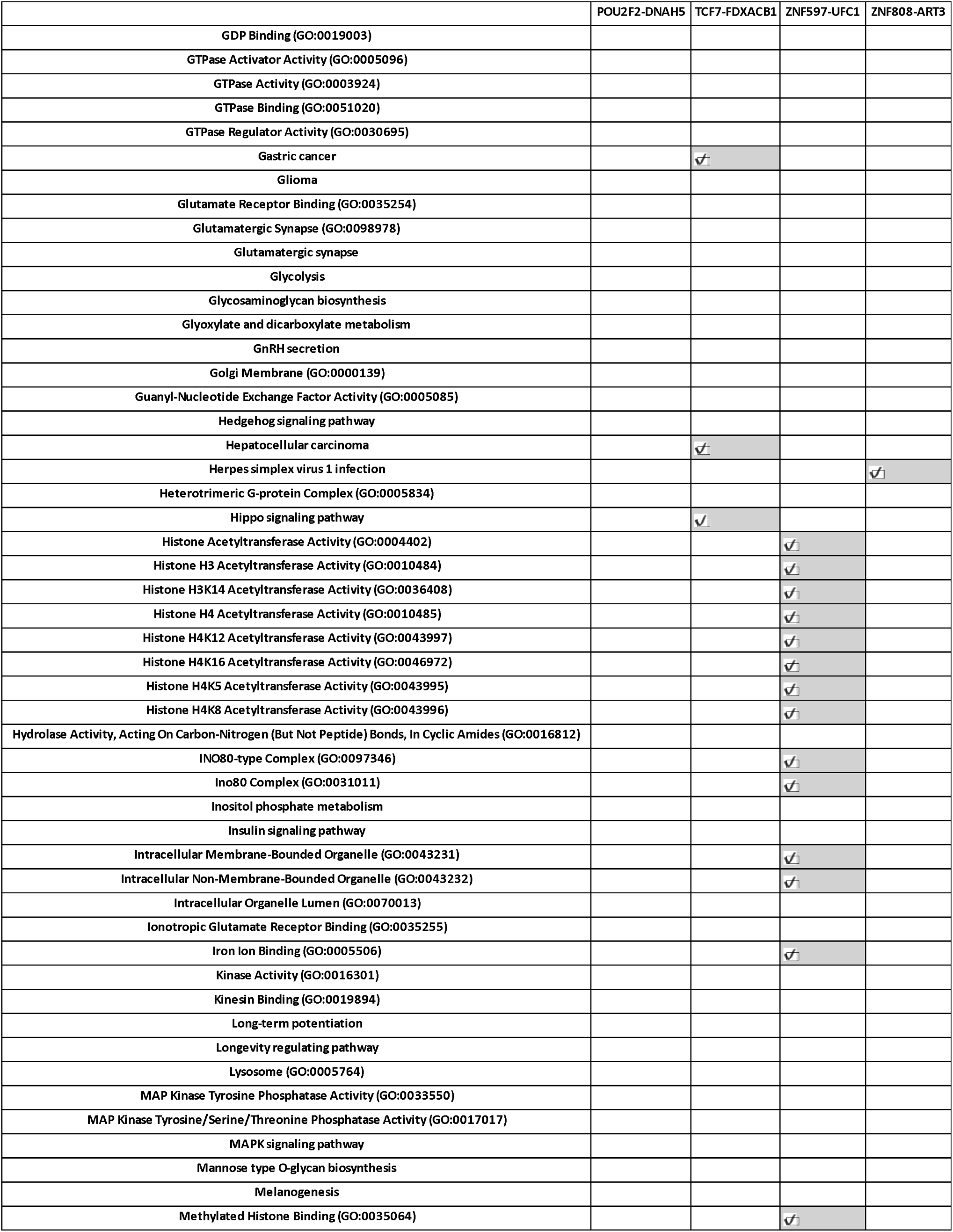
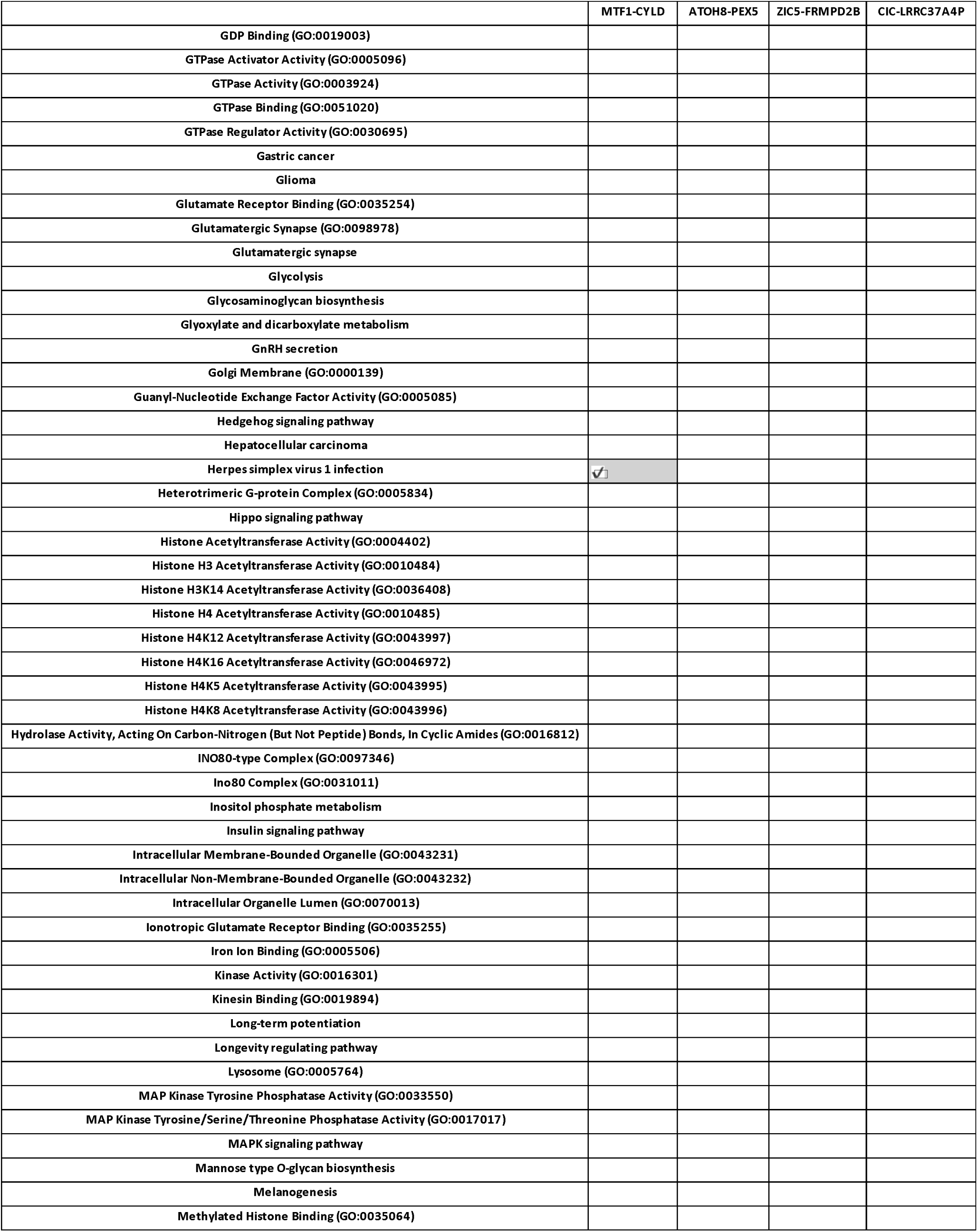
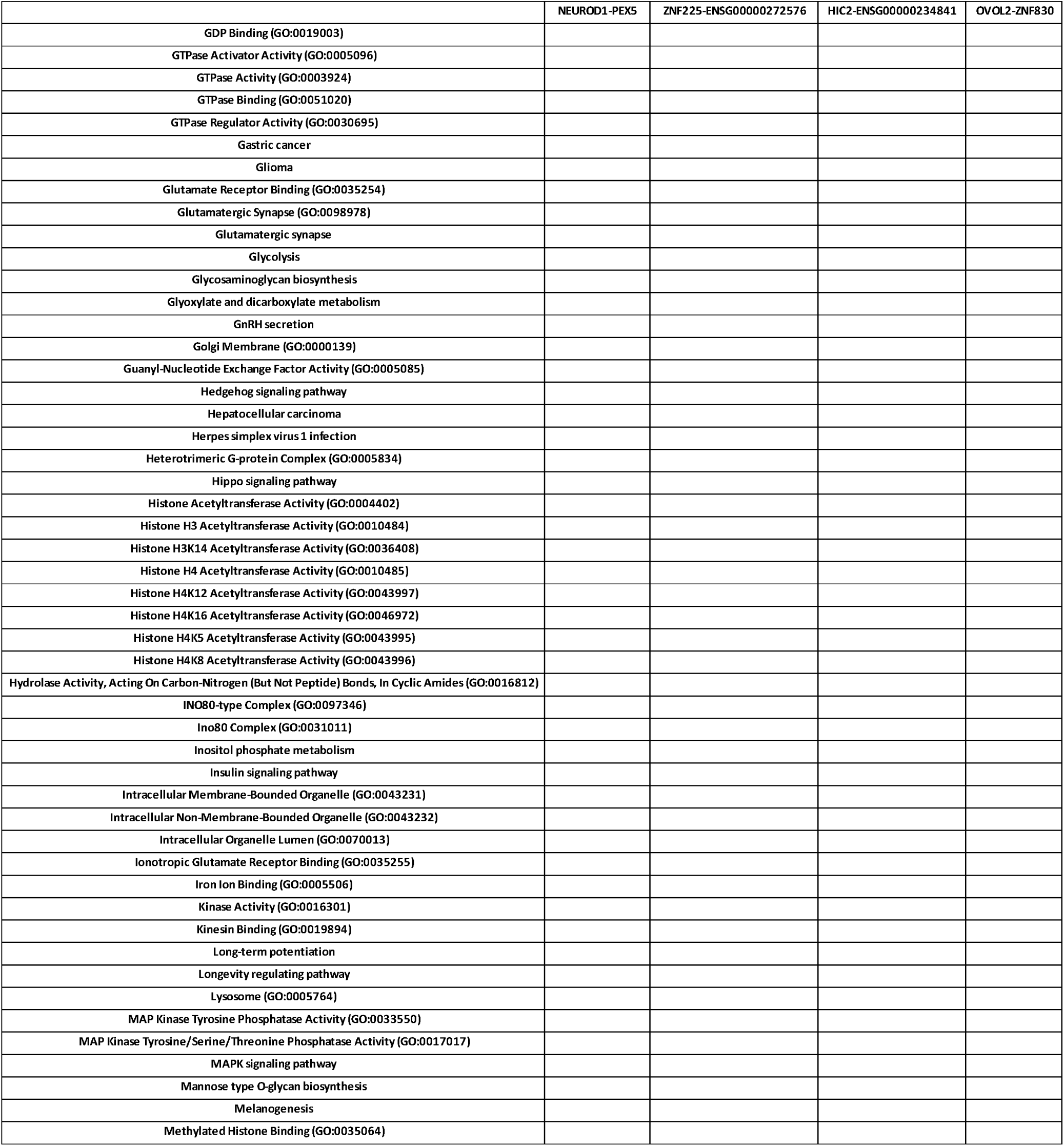
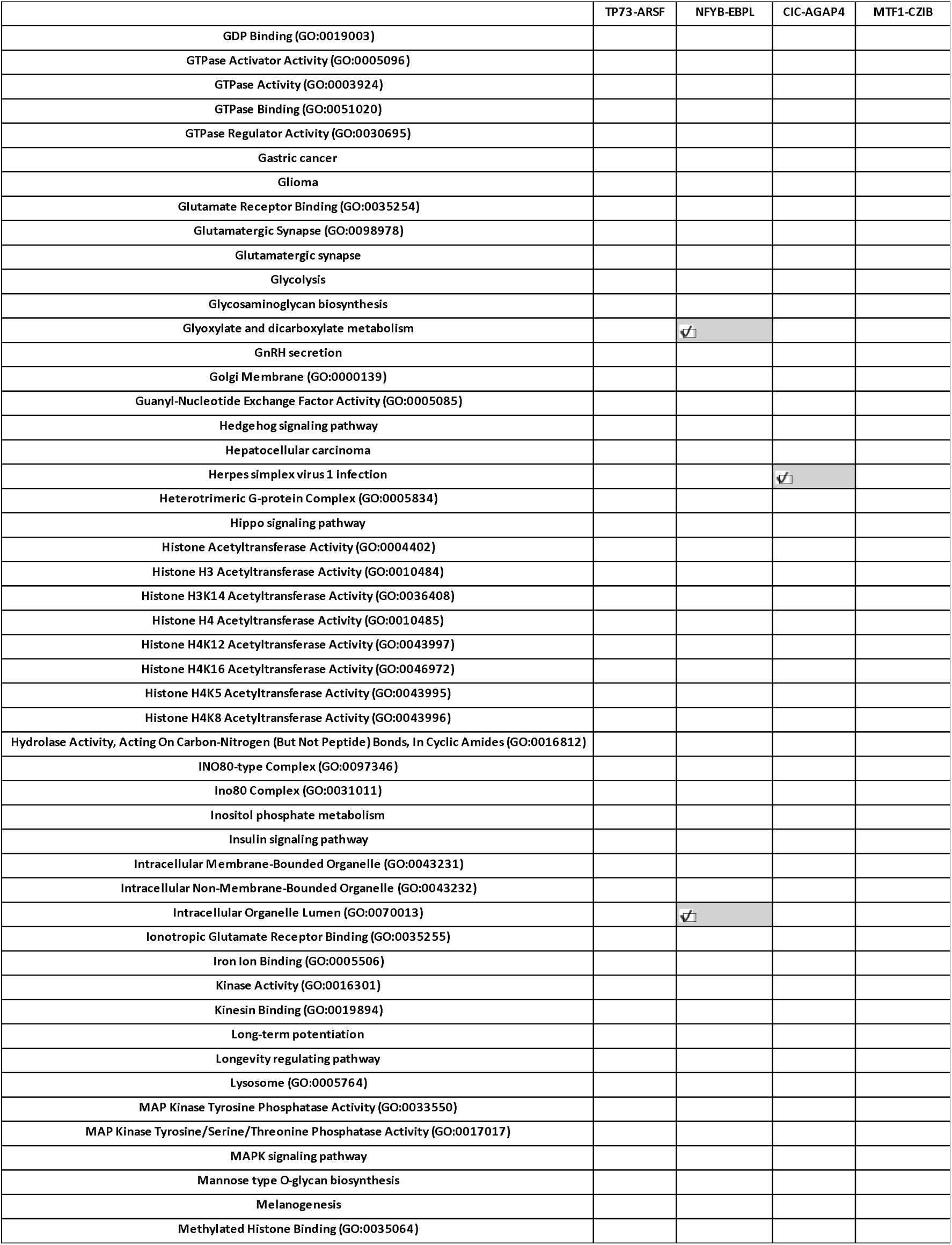
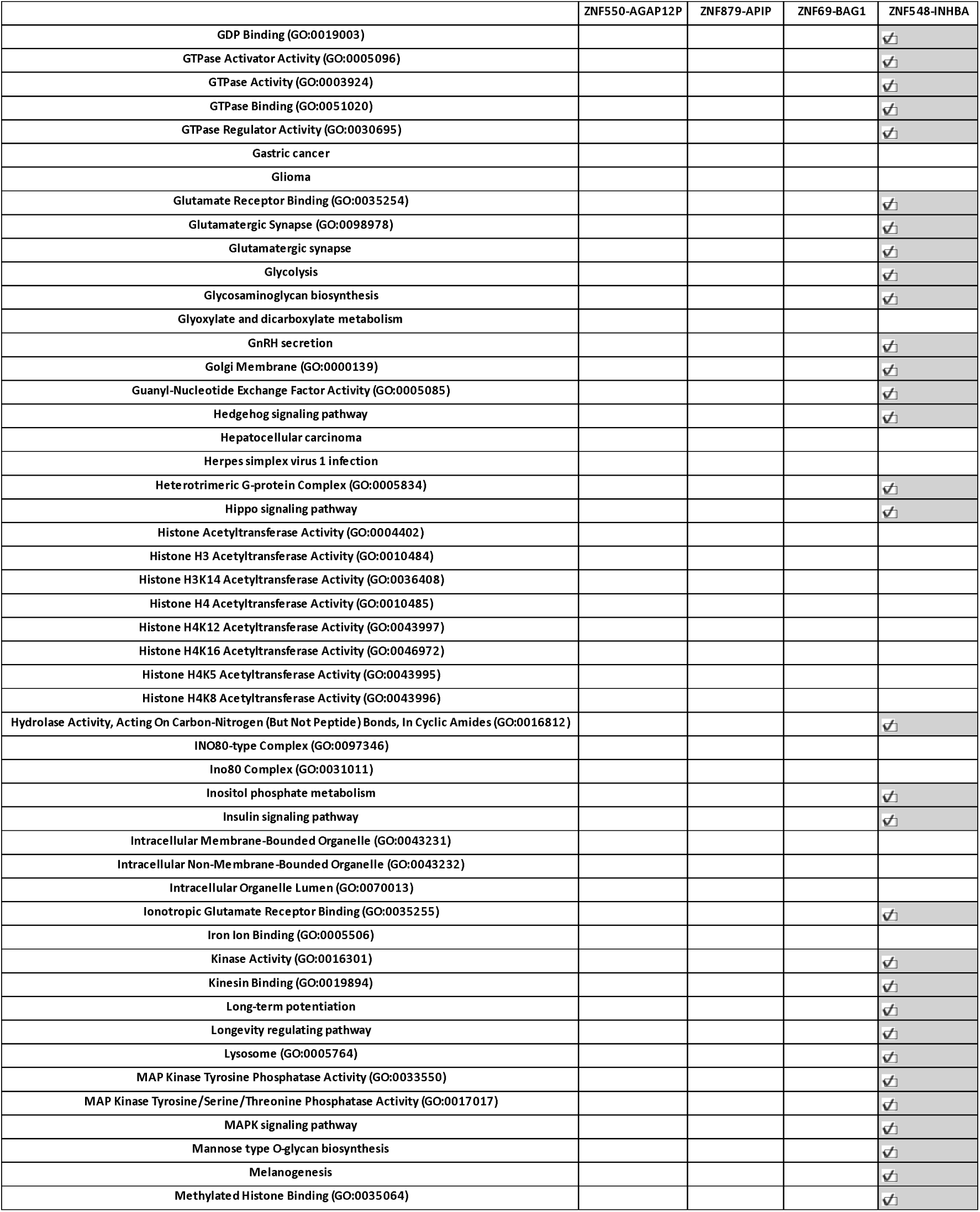
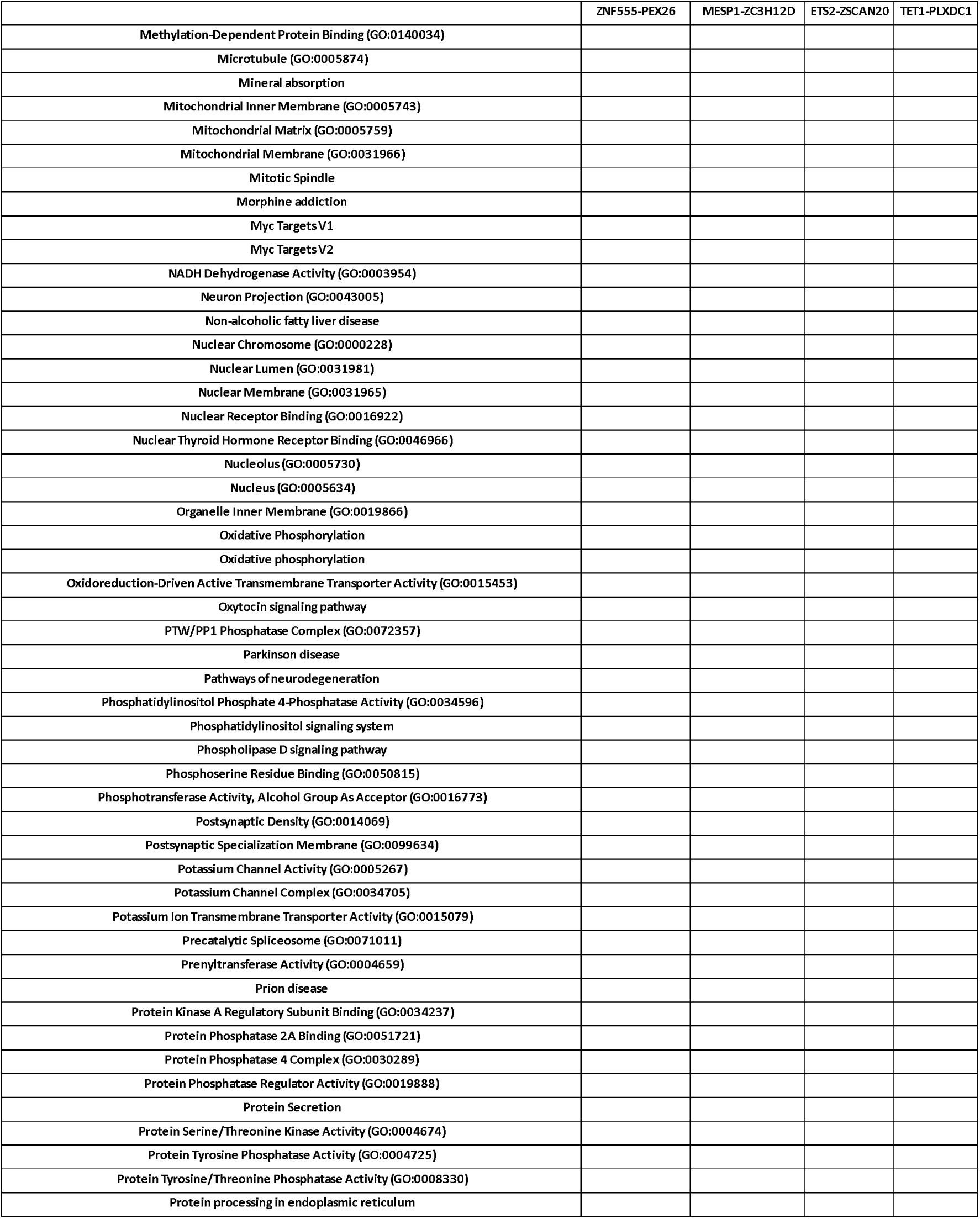
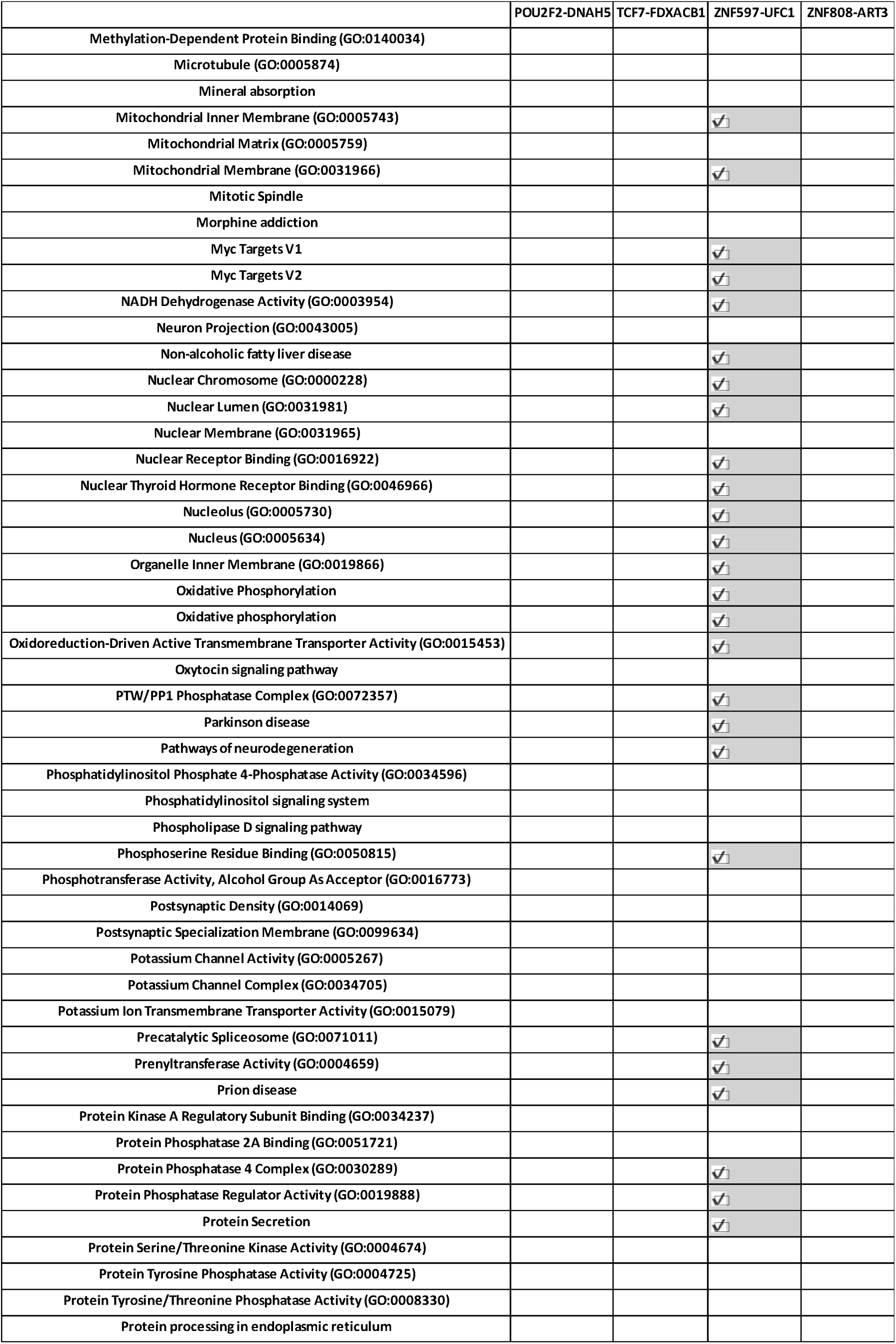
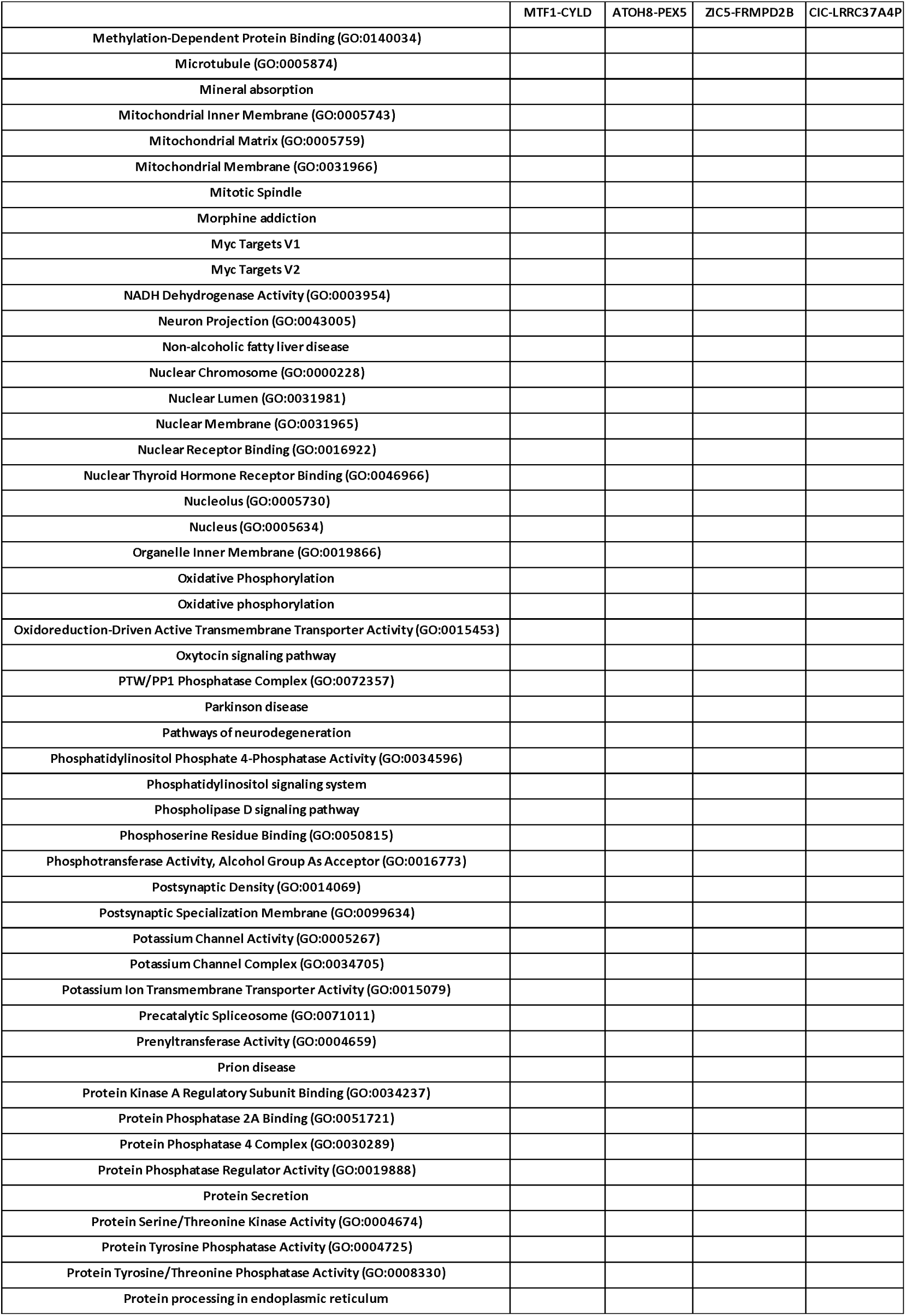
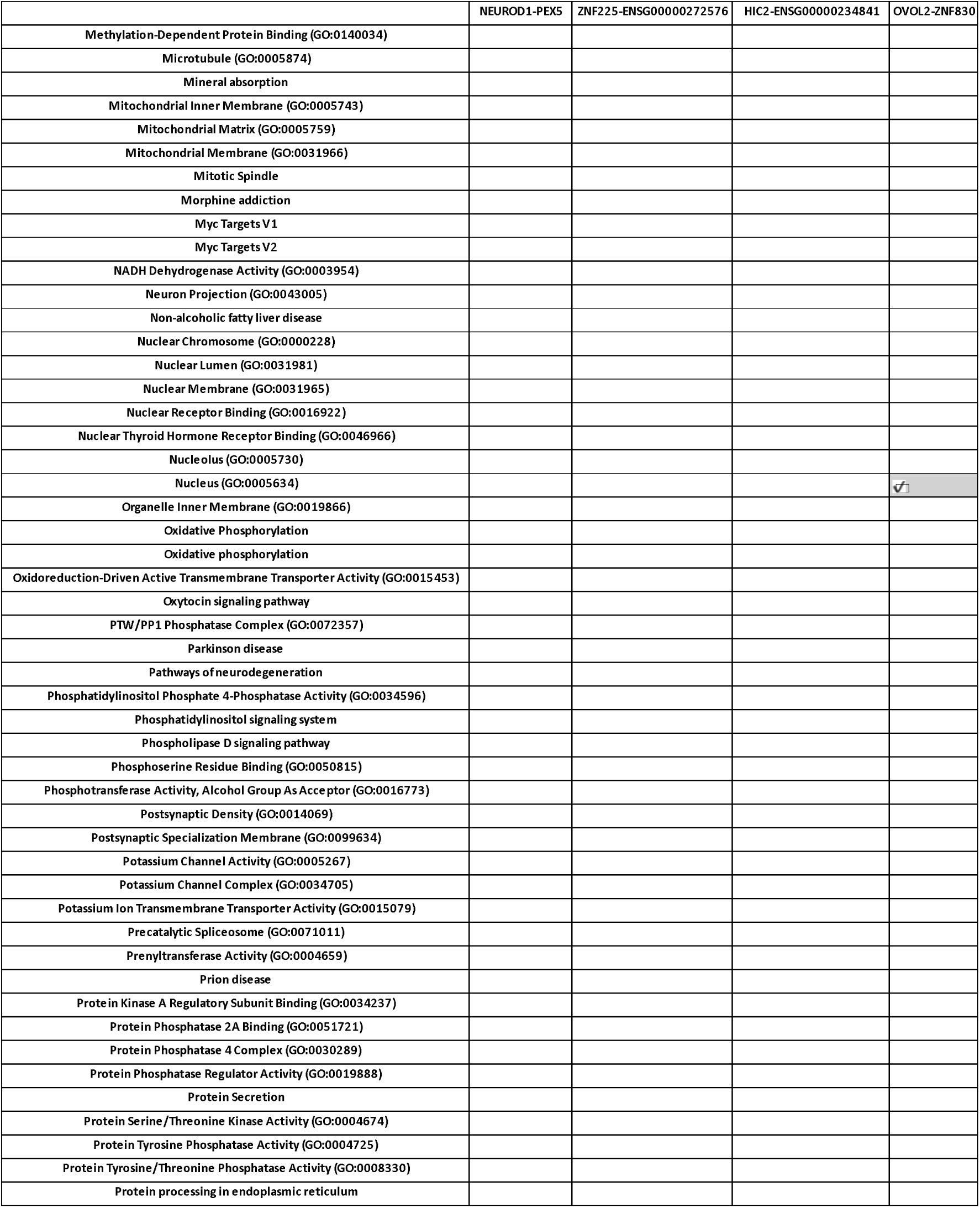
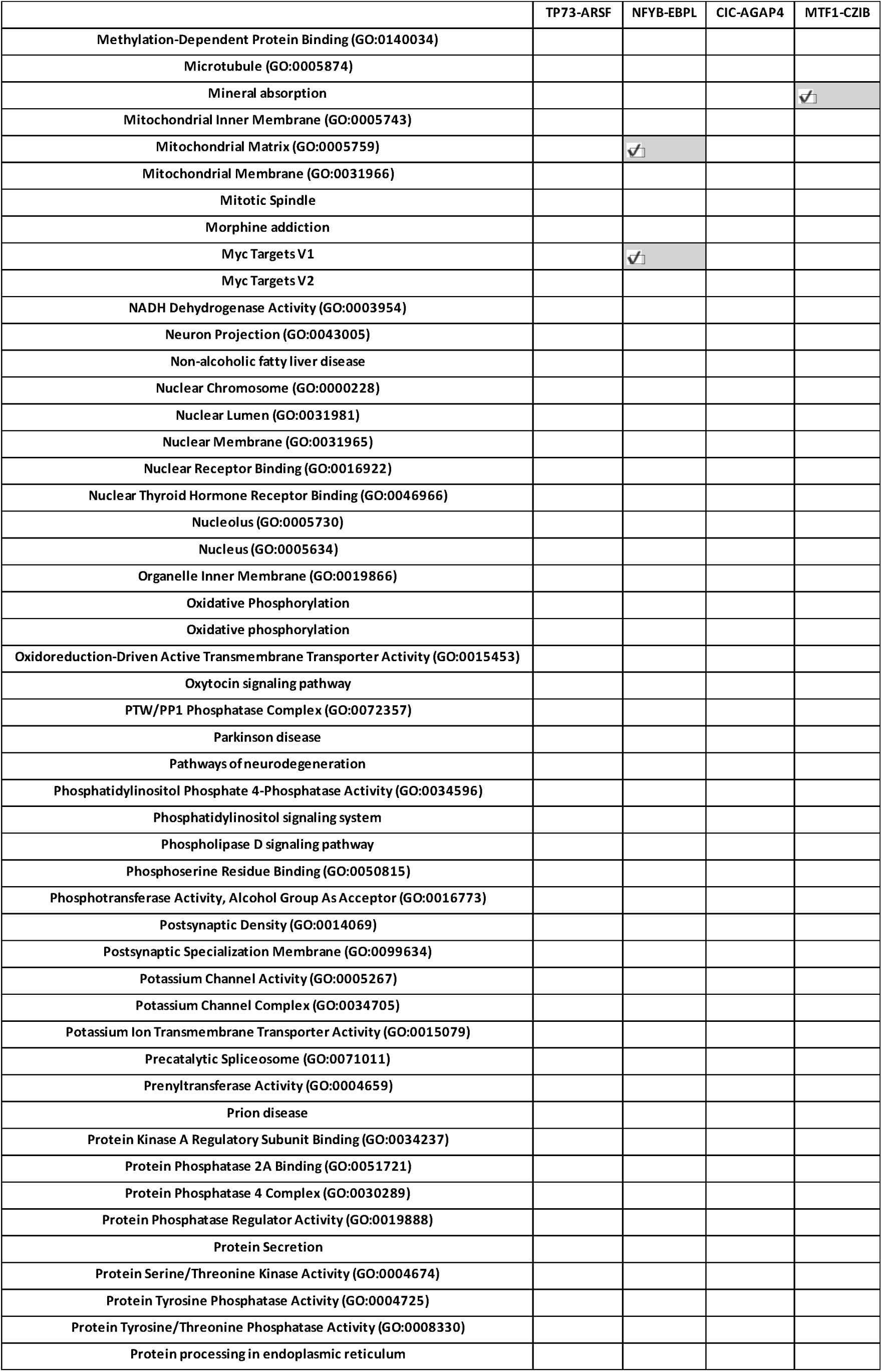
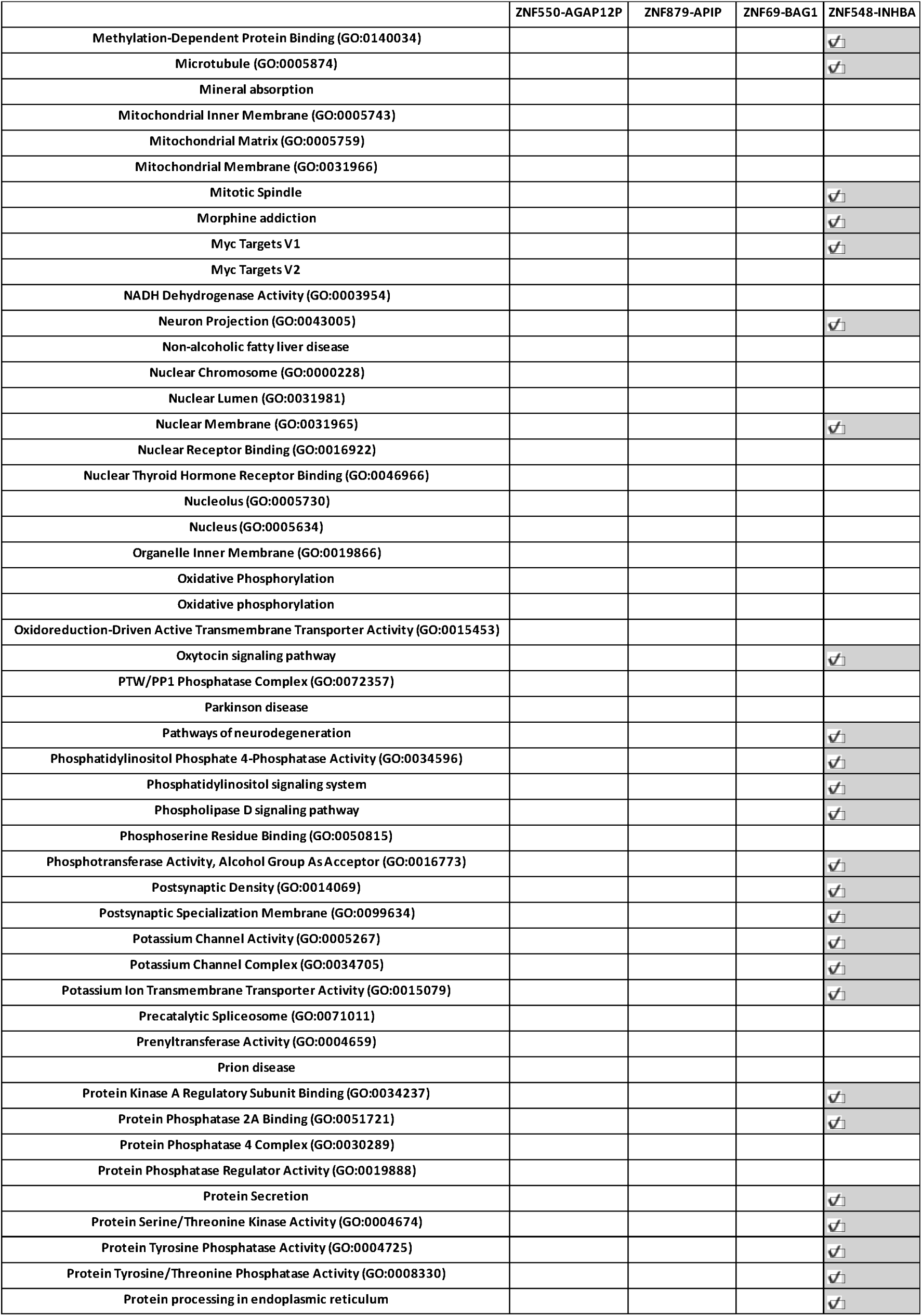
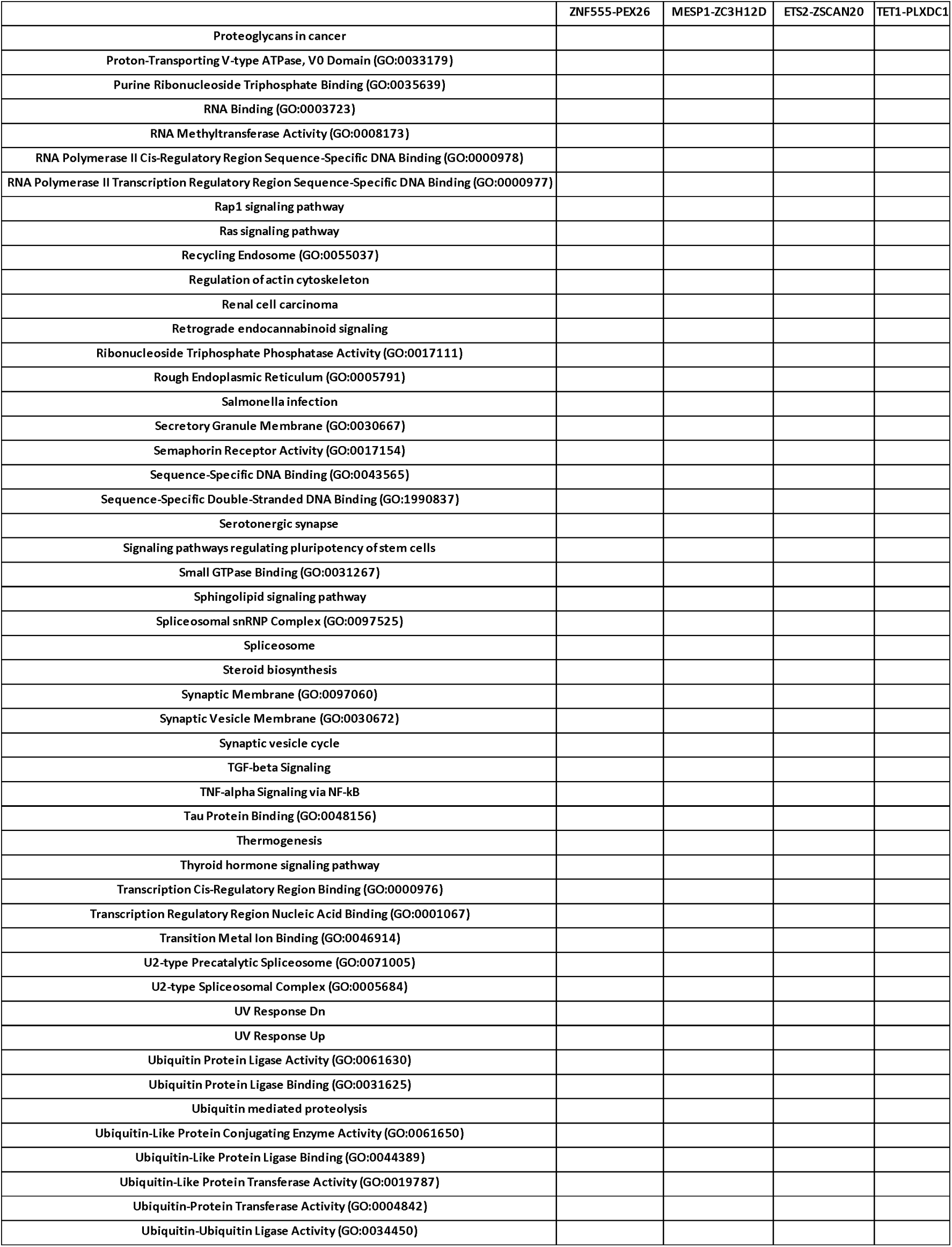
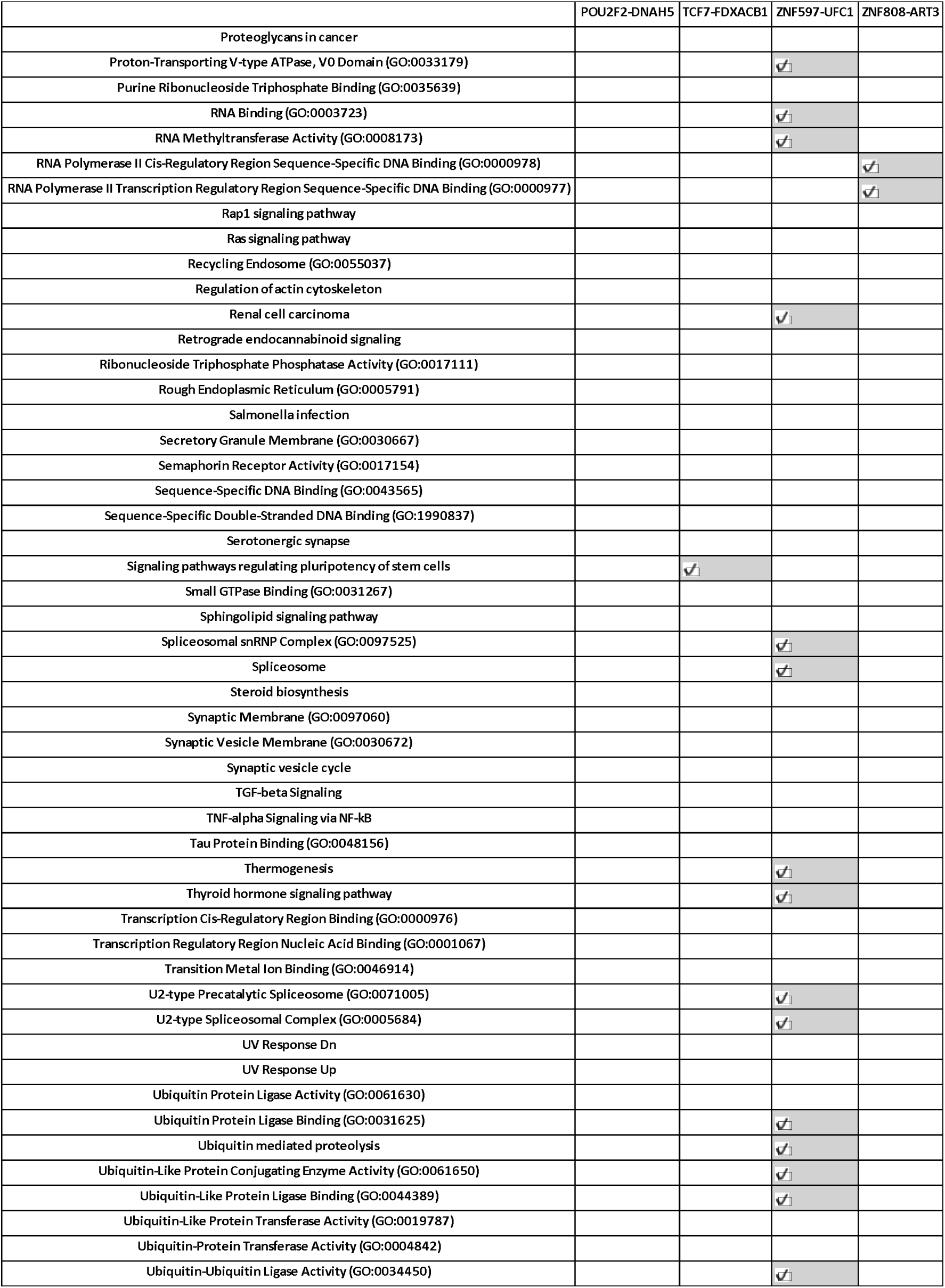
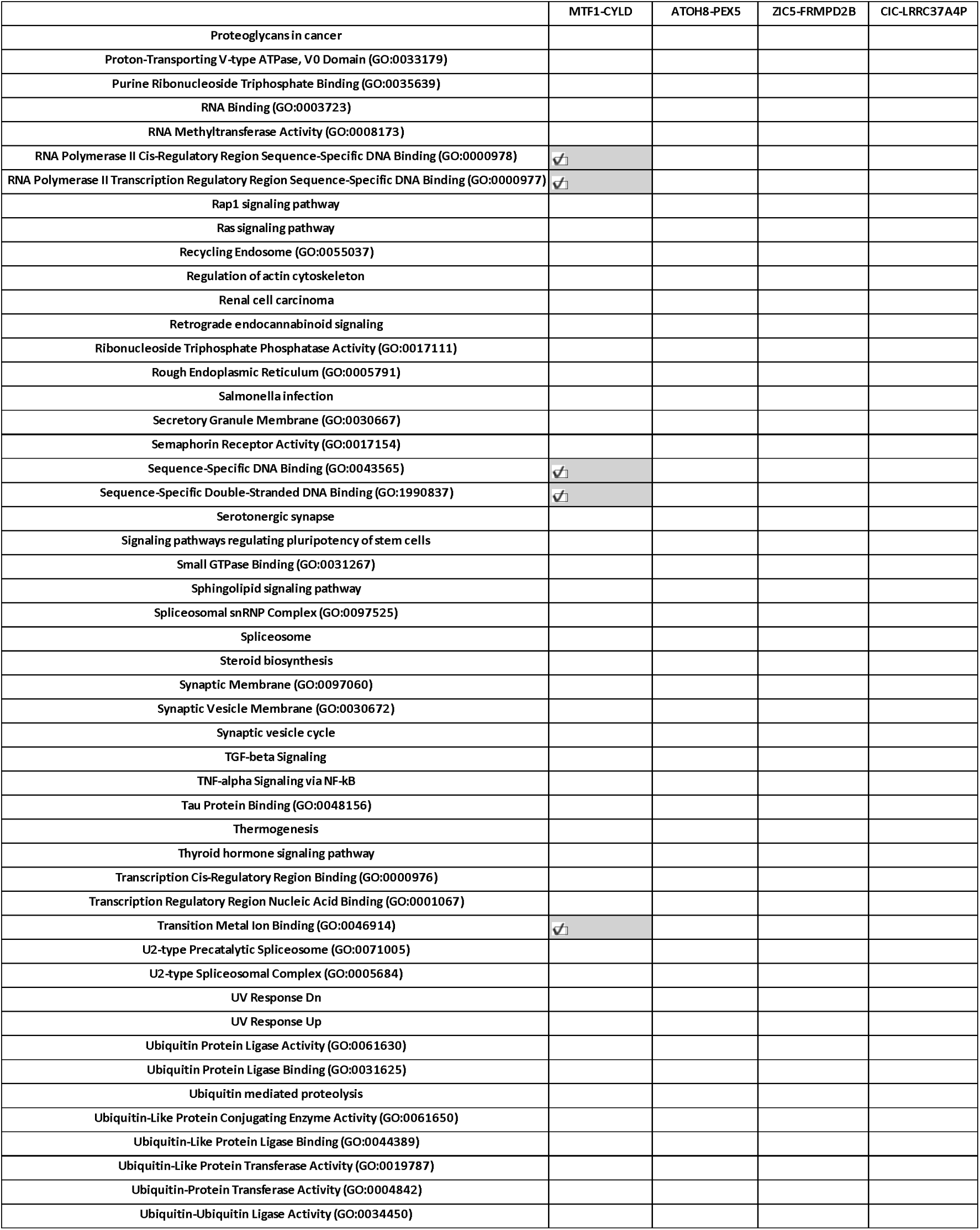
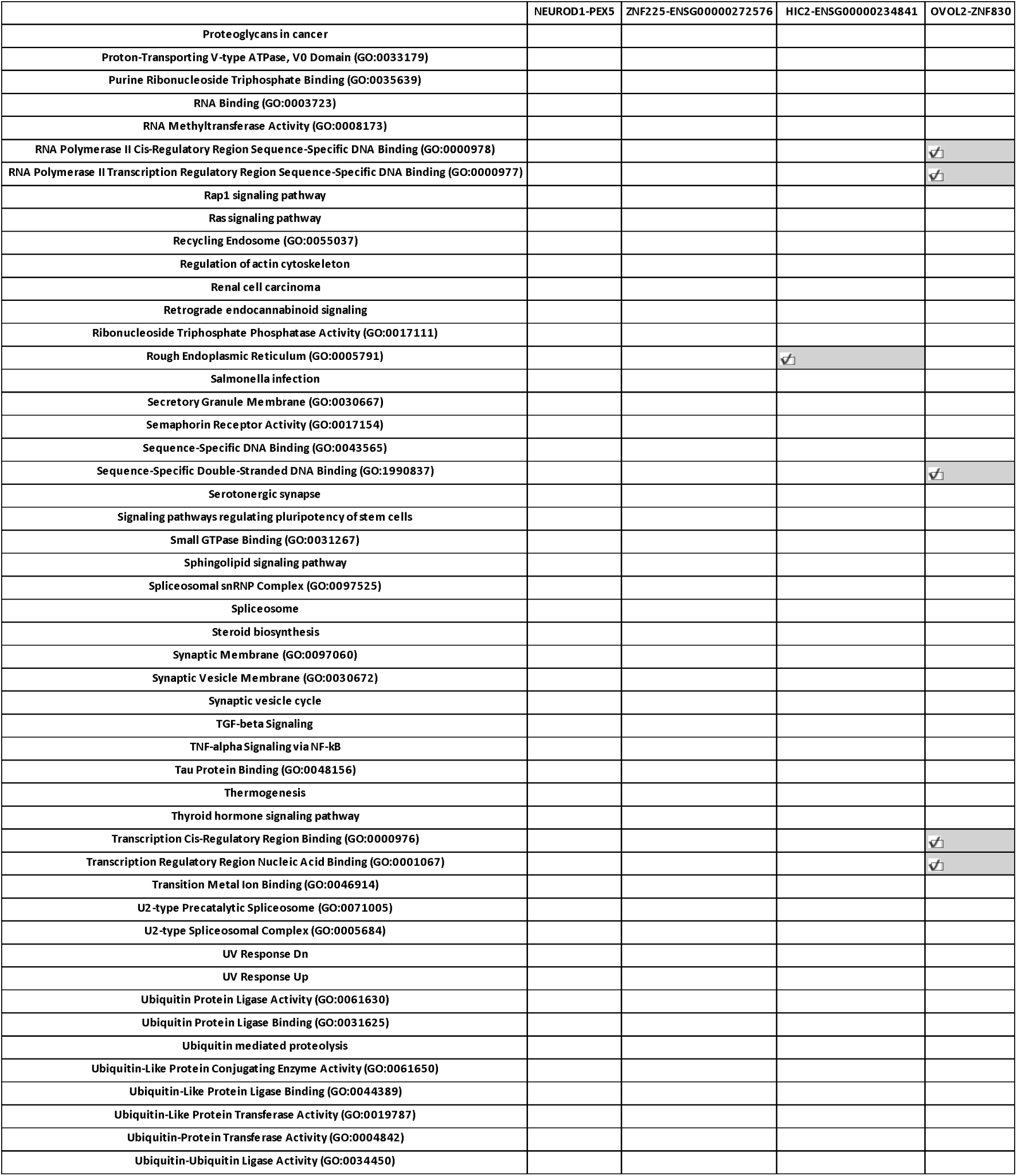
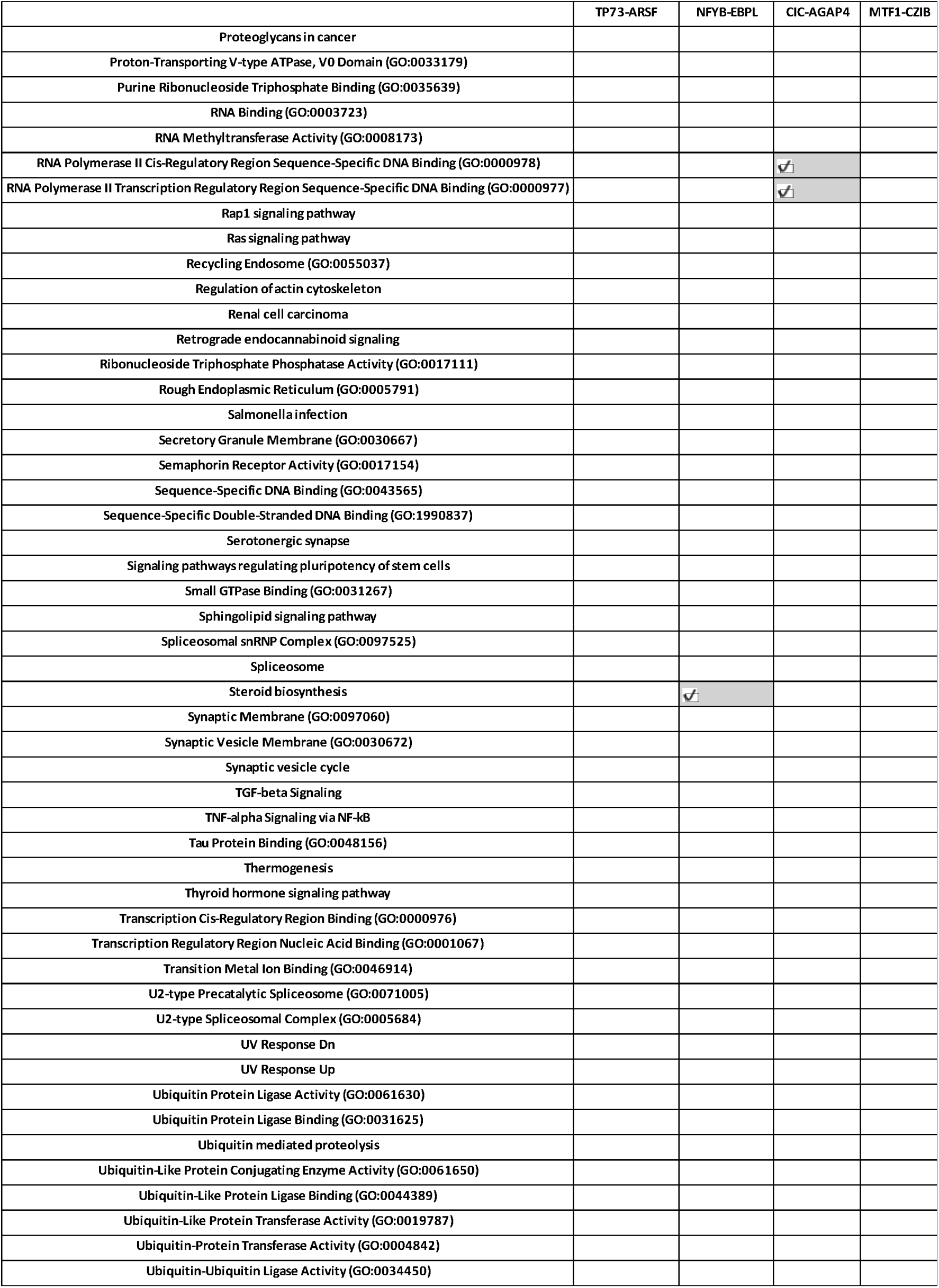
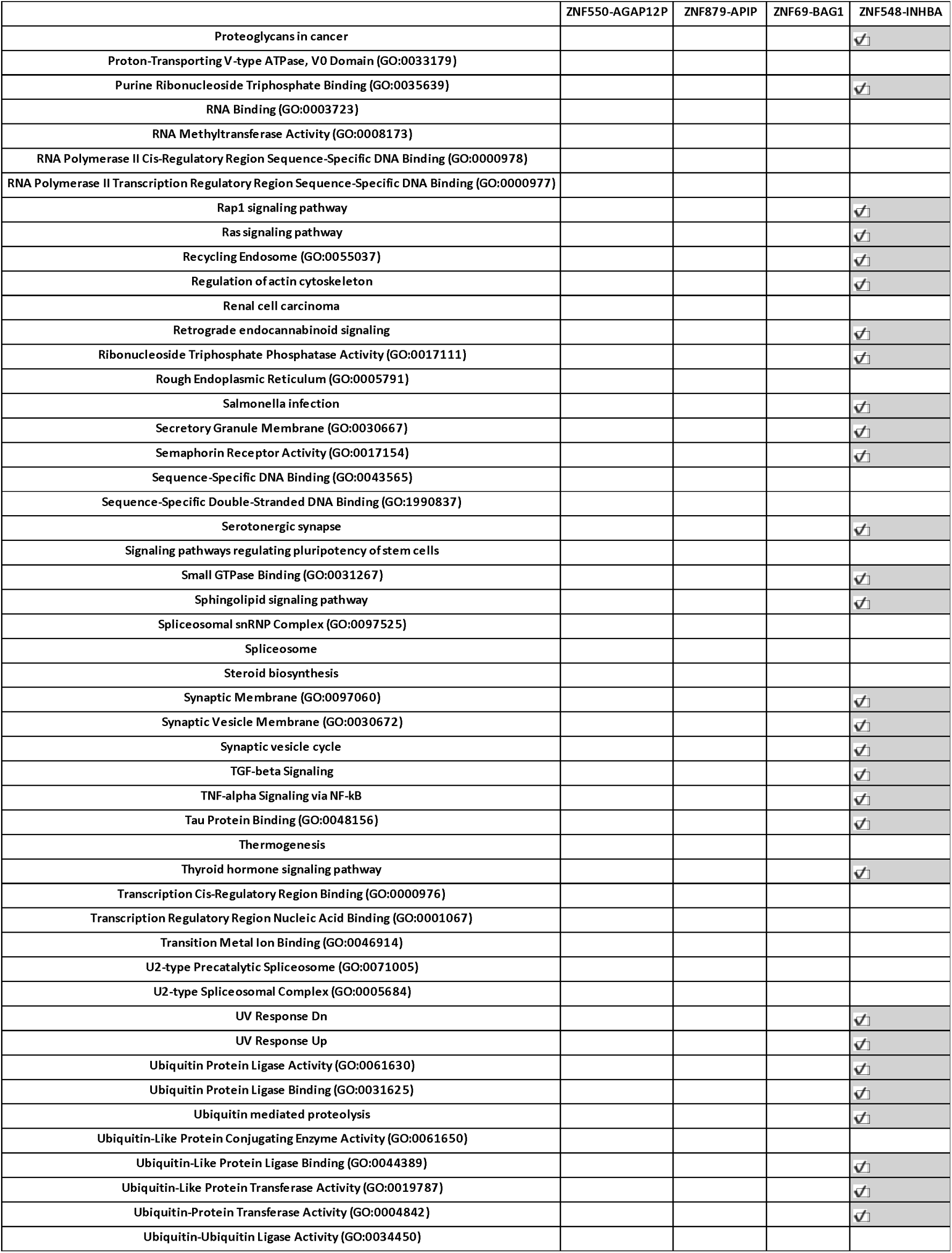
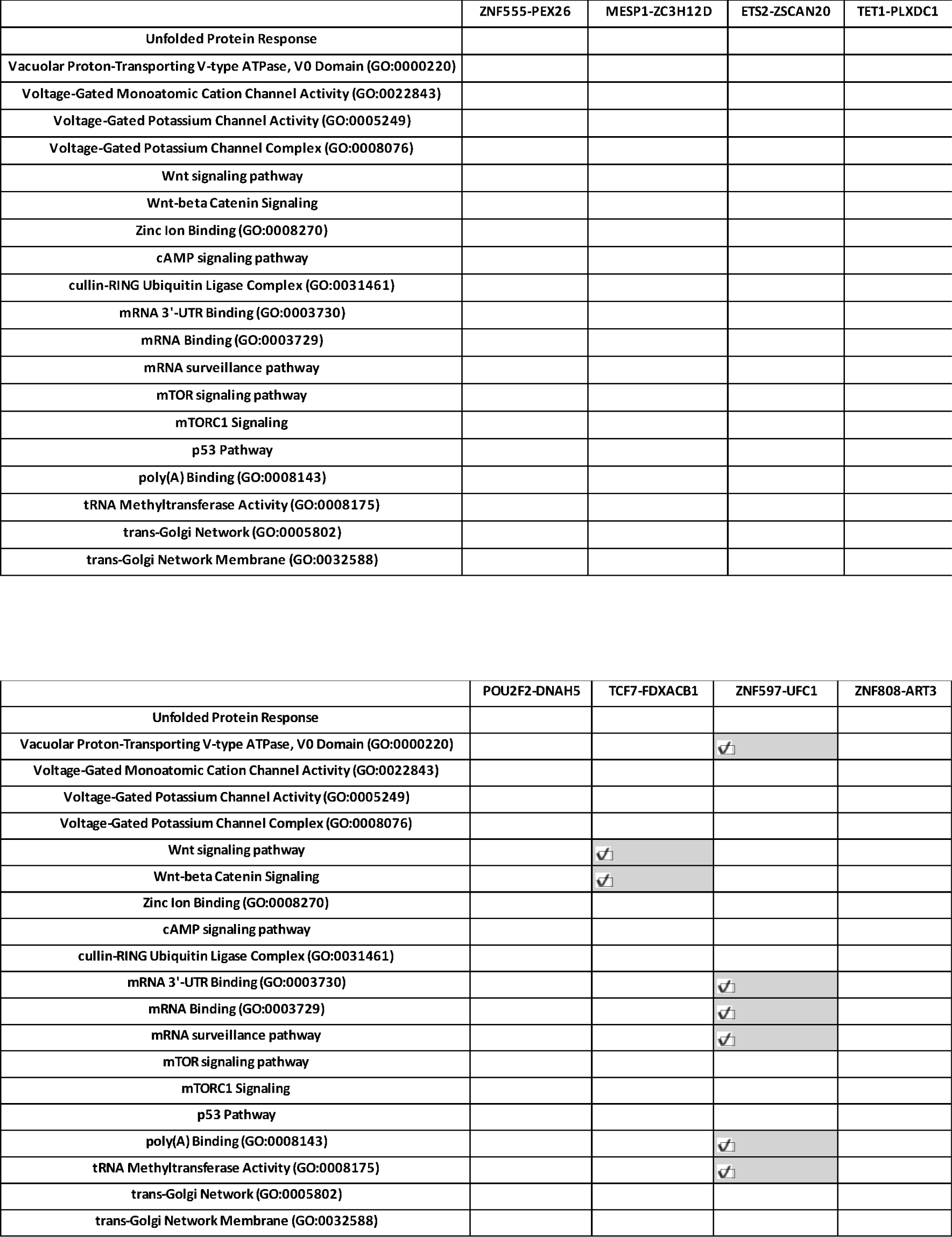
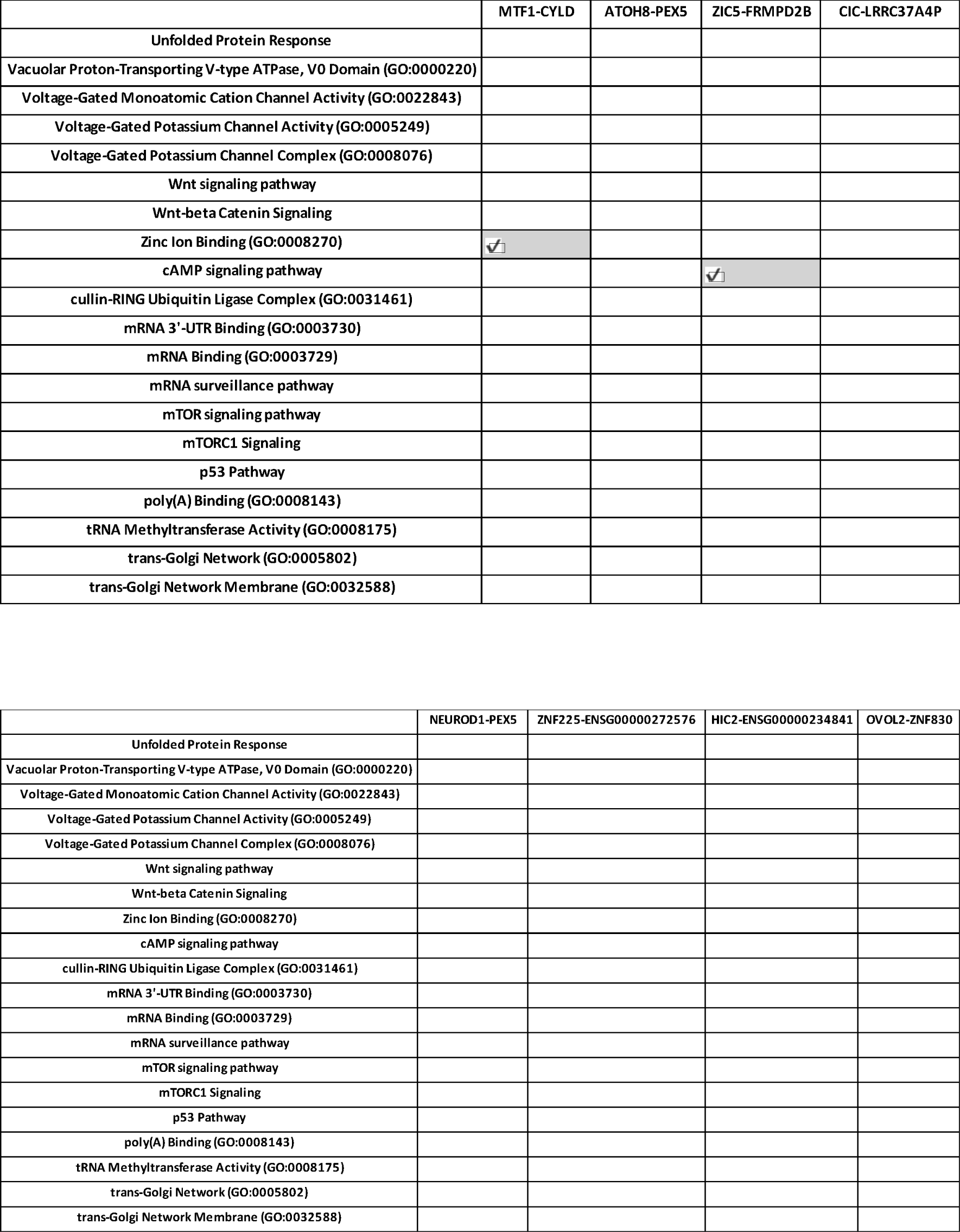
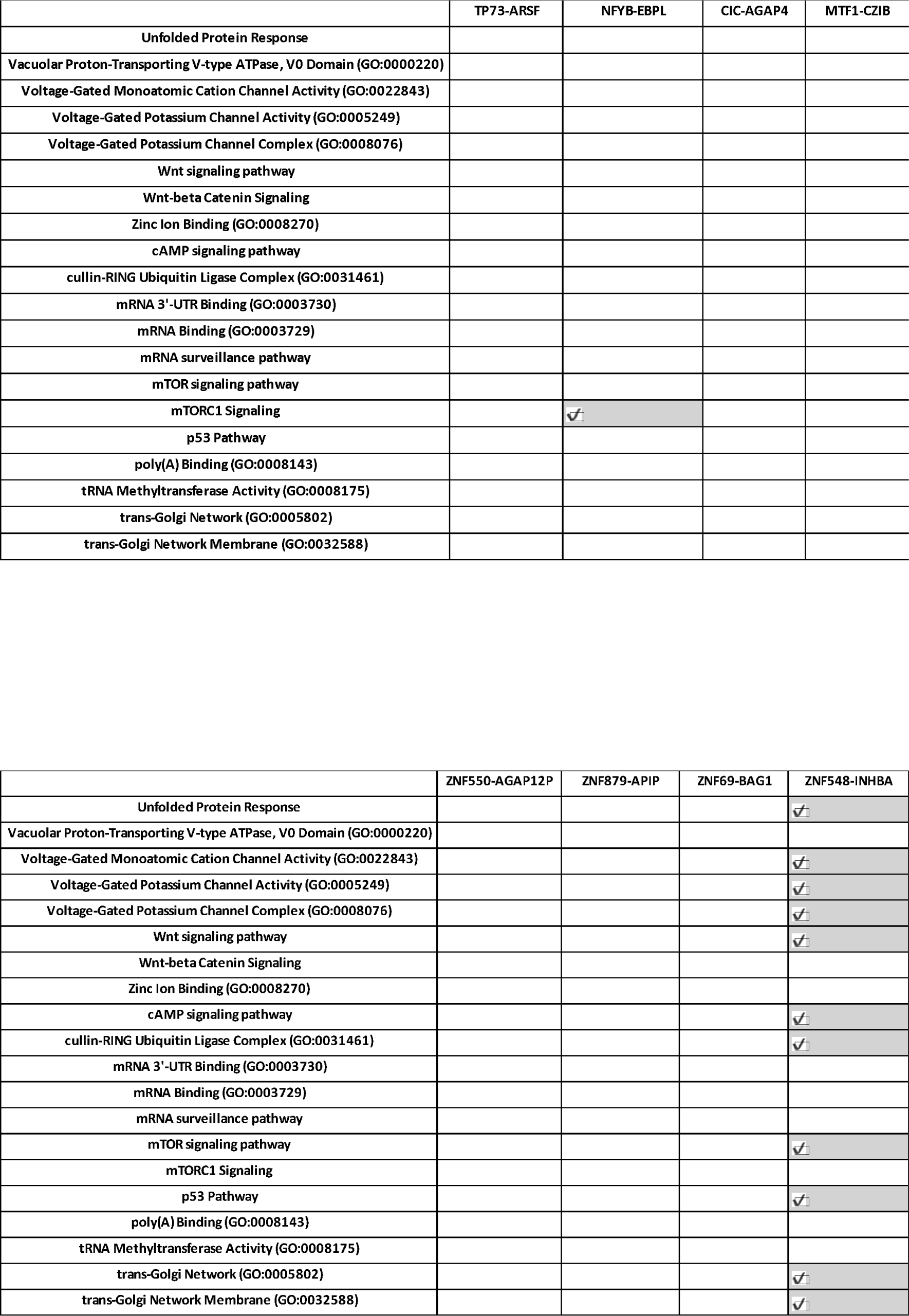
Enriched Pathways (p-adjusted value <= 0.05) corresponding to 22 key identified edges.

